# Online anxiety resources for Canadian youth: a systematic environmental scan

**DOI:** 10.1101/2022.08.09.22278279

**Authors:** Megan Pohl, Liza Bialy, Shannon D Scott, Lisa Hartling, Sarah A Elliott

## Abstract

**Introduction:** In a recent child health research priority setting exercise conducted in Alberta (CA), youth identified “mental health” as a priority topic. Specifically, youth were interested in understanding what the early signs and symptoms of anxiety were, and when they should seek help.

**Objective:** The objective of this study was to understand what information is currently available online for Canadian youth about the signs and symptoms of anxiety, what resources are available for self-assessment, and what are youth’s behaviors, experiences and information needs around seeking help for anxiety.

**Methods:** We conducted a systematic environmental scan of Internet resources and academic literature. Internet and literature search results (Information Sources) were screened by one reviewer and verified by another. Relevant information (e.g., self-assessment resource features and population characteristics such as age, presence of anxiety, and education) were then extracted and verified. Information Sources were categorized relating to the research concepts regarding; signs and symptoms, self-assessments, information needs and experiences. We complimented our environmental scan with youth consultations to understand how anxiety resources are perceived by youth, and what if anything, could be improved about the information they are receiving. Consultations were conducted over Zoom with three Canadian Youth Advisory Groups (2 provincial, 1 national) and took a semi-structured focus group format.

**Results:** A total of 99 Information Sources (62 addressing signs and symptoms, 18 self-assessment resources, and 19 reporting on information and help-seeking behaviors) met the inclusion criteria. The majority of Information Sources on signs and symptoms were webpage-based articles, and 36 (58%) specifically stated that they were targeting youth. 72% of anxiety self-assessment resources were provided by private institutions. The resources varied markedly in the post-assessment support provided to youth according to their source (i.e. private, academic, governmental). Regarding information and help-seeking preferences, three main themes were apparent and related to 1) obtaining in-person professional help, 2) searching for online help, and 3) stigma associated with seeking help for anxiety disorders. The Youth Group consultations identified several areas that need to be considered when developing resources for youth. The key considerations highlighted by youth across the consultations suggested resources needed to be; youth friendly, align with a credible institute (e.g. University, Health Institution), and provide useful resources post online assessment and tangible action items to support help seeking.

**Conclusion:** Awareness of the information and resources available to youth, coupled with an understanding of their help-seeking behaviors and information needs can help support the development and dissemination of appropriate knowledge translation tools around youth anxiety.

## Introduction

Anxiety disorders are the most common type of mental disorder (Phillips & Yu, 2021). Reports suggest 1 in 10 Canadians access health services for anxiety disorders each year and it has been “estimated that 2.4 million Canadians aged 15 years and older reported symptoms compatible with generalized anxiety disorder during their life time” (McRae et al., 2016a; Pelletier et al., 2017).

During the COVID-19 pandemic, Canadian youth have reported a significant decrease in their perceived mental health (Canada, 2020). It has been suggested that because of new public health measures and the loss of many activities that provide structure and meaning, the pandemic may be exacerbating youth anxiety (Courtney et al., 2020). Given that the prevalence of anxiety disorders is expected to increase as a result of the pandemic, supporting youths’ online access to quality mental health information is paramount (Courtney et al., 2020).

Generalized anxiety disorder (GAD) is characterized by excessive worry that is not specific to one trigger (e.g., social situations, open spaces, etc.) (Substance & Mental Health Services, 2016). According to the Diagnostic and Statistical Manual of Mental Disorders, 5^th^ Edition (DSM-V), anxiety and worry is difficult to control and is associated with restlessness, becoming easily fatigued, difficulty concentrating, irritability, muscle tension, and sleep disturbances (Substance & Mental Health Services, 2016). The early identification of GAD is often challenging, as many individuals consult a health care provider (HCP) for somatic symptoms such as fatigue, trouble sleeping, headaches, gastrointestinal symptoms or symptoms related to comorbidities, and not for anxiety or worry (Hoge et al., 2012; Nutt et al., 2006).

Untreated, youth anxiety disorders often persist into adulthood and are associated with multiple maladaptive outcomes such as suicidal tendencies, depression, poor educational outcomes, and substance abuse (Kendall et al., 2004; Pine et al., 1998). Treatment is often indicated when an individual experiences distress resulting from the disorder or additional complications such as suicidal ideation (Bandelow et al., 2017).

There is increasing evidence suggesting that intervention during the early stages of mental health disorders may help reduce the severity and/or the persistence of the initial or primary disorder and prevent secondary disorders (de Girolamo et al., 2012). Despite the pre-adult onset of most mental disorders, young people are less likely than any other age group to access mental health services for a number of reasons such as stigma, reduced mental health literacy, poor access to appropriate services and inadequate health system structures (Rickwood et al., 2007). In 2019, a systematic review identified ‘text-based internet searching’ as the most common help-seeking approach used by young people to find mental health information (Pretorius et al., 2019). However, little is known about what online resources are available to youth regarding anxiety, as well as their online information needs.

Furthermore, in a recent child health research priority setting initiative conducted in Alberta, Canada, youth (aged 15-24 years), identified “mental health” as a priority topic (Elliott, 2021). Within that, the most highly ranked question by youth was, “What are the signs and symptoms of anxiety and when should an individual seek help?”. As youth search for mental health information online, it is important to identify and categorize what is currently available to youth online, as well as their information needs and preferences for help seeking.

To identify information that addresses this priority topic, we conducted a systematic environmental scan to provide a comprehensive description of what information exists around three key questions:

1. What information is available online for Canadian youth about the signs and symptoms of anxiety?
2. What self-assessment resources for anxiety are available online for Canadian youth?
3. What are youth’s experiences and information needs around seeking help for anxiety?

After conducting the environmental scan, we consulted youth collaborators to understand how the resources were perceived by youth, and what could be improved to enhance their use.

## Methods

We conducted a systematic environmental scan of Internet-based resources as well as searching the primary literature. While there is no set definition or guidance for conducting an environmental scan, it is a widely used method for gathering information about current and emerging issues through a systematic search of websites and other sources (Charlton et al., 2019; Graham et al., 2008; Wilburn et al., 2016). This methodology was chosen as we anticipated needing to scan various sources to answer these diverse questions. We report our findings (whenever possible) according to the Preferred Reporting Items for Systematic reviews and Meta-Analyses extension for Scoping Reviews (PRISMA-ScR) checklist (Tricco et al., 2018). For this systematic environmental scan three main sources were searched: primary/scientific literature, scholarly literature via Google Scholar, and Internet websites via Google search engine. The term “Information Source” was used to describe any type of primary literature, Google, and Internet sources retrieved.

### Search Methods

#### Primary Literature

Utilizing concepts for each of the three key questions, in August 2020 a research librarian carried out searches in Ovid Medline, Ovid Embase, Ovid PsycINFO, CINAHL via EBSCOhost, and Wiley Cochrane Library databases: Cochrane Database of Systematic Reviews, Database of Abstracts of Reviews of Effects and the Health Technology Assessment Database. These search results were limited to publications in English since 2015 to ensure the most recent literature was included and reflected current internet usage amongst youth. To ensure a comprehensive coverage of all Information Sources we reviewed the reference lists of relevant studies found through the search strategy.

#### Google Scholar and Internet Search

Google Scholar and Google were searched, utilizing the following key concepts: youth, anxiety, App (an application downloaded by a user to a mobile device), signs/symptoms, help-seeking, assessment tool, knowledge translation, and information needs. For each search term the links on the first 10 pages retrieved by Google and Google Scholar were assessed.

The complete search strategy is available in Appendix 1. Search results were exported to EndNote X7 (Clarivate Analytics) and duplicates removed prior to screening in Microsoft Office Excel (v. 2016; Microsoft, Redmond, Washington, USA).

### Screening

#### Inclusion and Exclusion

The following eligibility criteria (Appendix 2) were applied to all Information Sources (academic articles, websites, resources etc.): (a) published in English, (b) from a developed country (Prospects, 2014), (c) published or created from 2015 onwards, (d) included or applicable to youth between 15 and 24 years of age ((UNDESA), 2008). There were no restrictions on study design or format of Information Source.

For this review, Information Sources such as ‘self-assessment resources’ were included if a series of questions was presented relating to the user’s anxiety level, and was intended for self-administration by the youth/general public/non-health professionals. Information Sources for signs and symptoms were included if a listing or description was provided and geared towards youth with anxiety. To be included as a help-seeking Information Source there needed to be information about help-seeking behaviours, suggestions on where to seek help or information needs desired by youth.

Information Sources were excluded if reporting on anxiety was as a result of managing chronic illness, substance abuse, previous suicide attempts, eating disorders, bullying/victimization, divorce, trauma, racial discrimination, food/home insecurity during childhood or other anxiety related disorders (e.g., obsessive-compulsive, panic, or post-traumatic stress disorders, social phobia, social anxiety disorder, needle/pain anxiety, etc.).

Based on a priori eligibility criteria one experienced reviewer screened the titles and abstracts of all Information Sources and classified as “include/unsure” or “exclude”. A second reviewer assessed all categorized as “exclude” to confirm decisions. For full-text screening the same process was used to review all excluded sources in duplicate. For the Internet search, websites were only excluded if they were clearly not related to anxiety by one reviewer.

#### Data Collection

One reviewer extracted data from each included Information Source into Microsoft Office Excel with another reviewer verifying a 10% random sample for each key question. The following data were extracted: Information Source type (website, app, academic article), country of publication, institution type, study design (if applicable), population characteristics (or target audience) such as age, gender, presence of anxiety, and education. Definitions specific to the key questions and data extraction can be found in Appendix 3.

#### Data Analysis

Descriptive summaries and graphical formats were used to synthesize information regarding Information Source characteristics. Where appropriate, conventional content analysis (Hsieh & Shannon, 2005) was applied to identify patterns in identifying signs/symptoms, help-seeking behaviours, information needs, and self-assessment resources and App functionality.

#### Collaborator Engagement

For contextualization of the environmental scan results, we engaged our youth collaborators (1 national and 2 provincial Youth Advisory Groups) who identified several areas that need to be considered when developing resources for youth. Discussions with the groups regarding the readability, usefulness and youth friendliness of the identified online anxiety resources helped direct considerations put forth for future resource development.

## Results

We identified a total of 12,874 Information Sources from the primary literature, Google Scholar and Internet search via Google, combined. After title and abstract screening 446 full-text Information Sources were reviewed and a total of 99 Information Sources were included (Figure 1).

**Figure 1:**
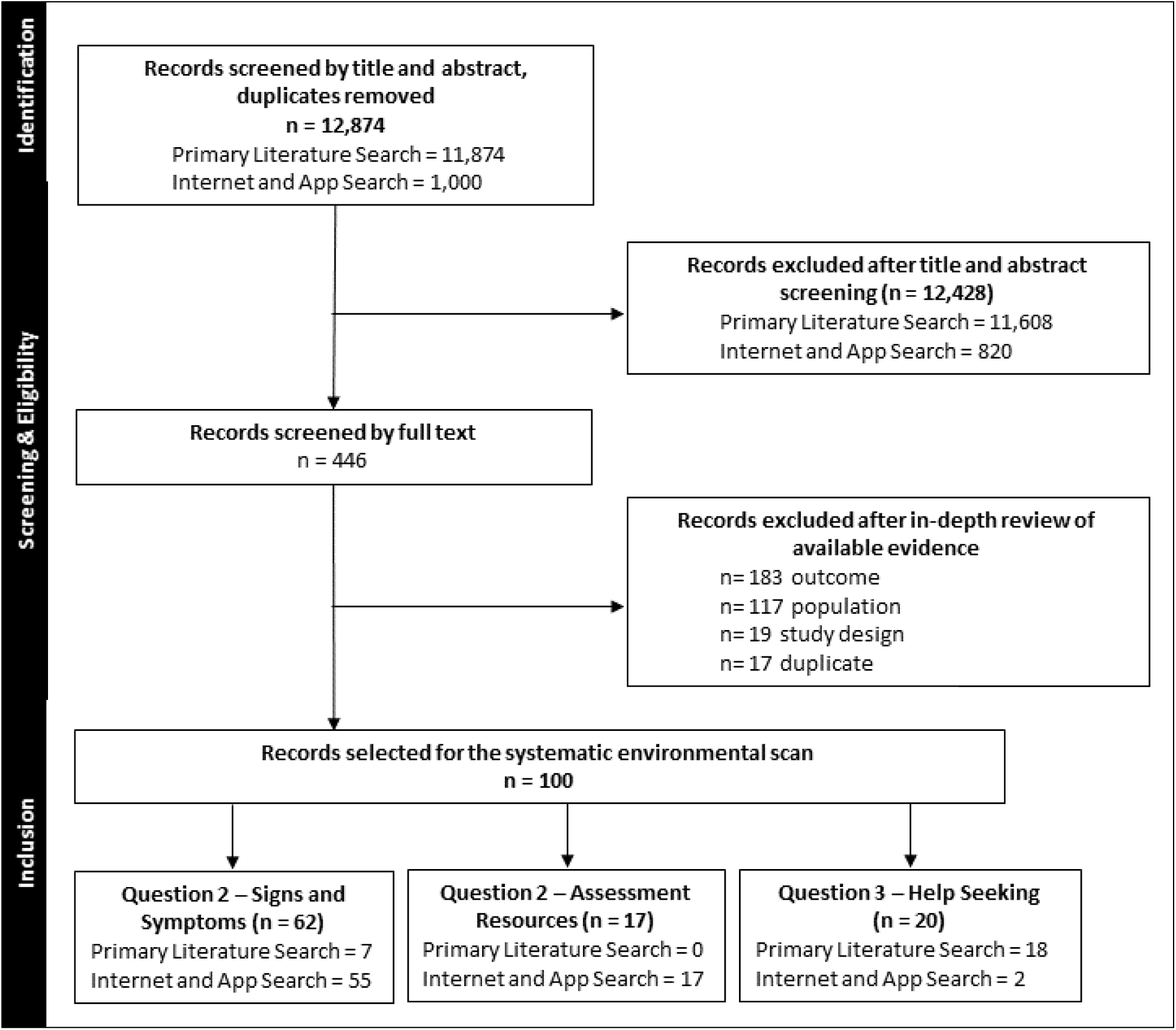
Flow diagram of search strategy

### What information is available to youth about the signs and symptoms of anxiety?

#### Characterizing signs and symptoms

Sixty-two Information Sources addressing the signs and symptoms of anxiety were identified (Table 1). The majority of Information Sources were webpage-based articles (n=53, 85%). Other Information Sources included narrative reviews (n=4, 6%), cross-sectional studies (n=3, 5%), a single prospective cohort (n=1, 2%) and a qualitative study (n=1, 2%). Most Information Sources originated from the USA (n=25, 40%) and Canada (n=21, 34%), followed by Australia (n=6, 10%) and the United Kingdom (n=5, 8%). Information Sources were developed by private companies/individuals (n=20, 32%), government agencies (n=18, 29%), private foundations/charities (n=13, 21%), and academic institutions (n=11, 18%).

**Table 1:**
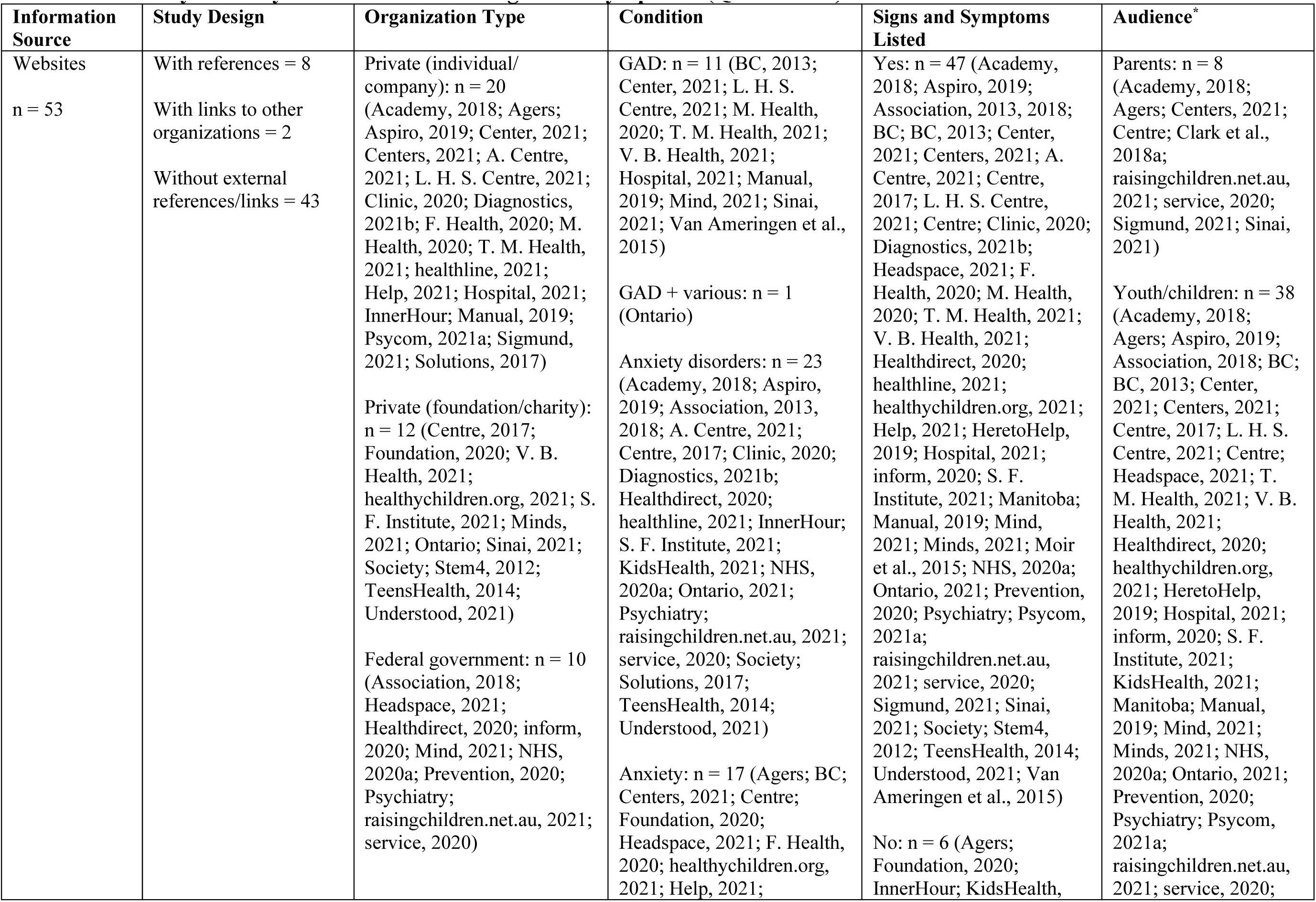
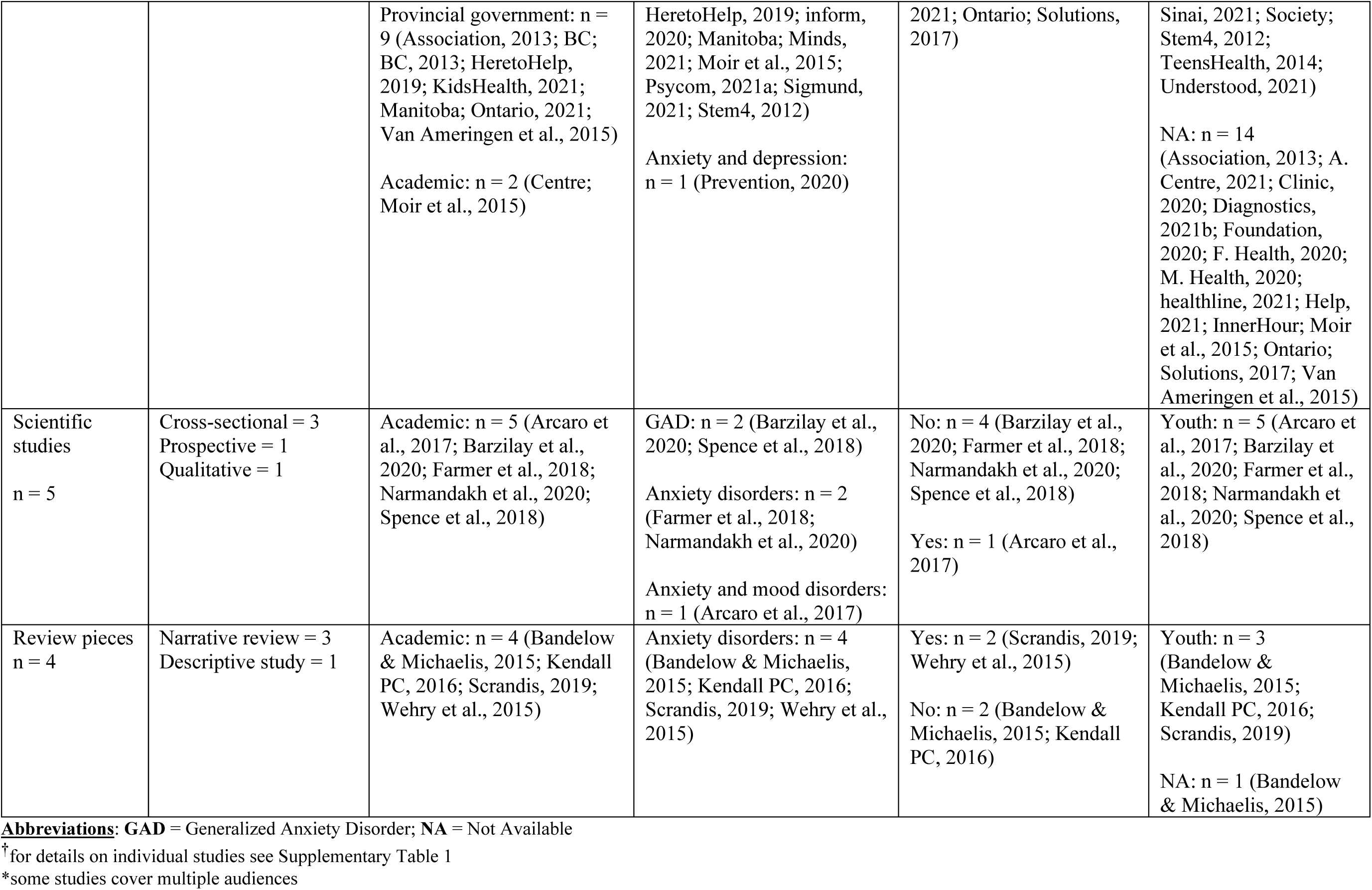
Summary of Study Characteristics for Signs and Symptoms (Question #1)^†^

| Information Source | Study Design | Organization Type | Condition | Signs and Symptoms Listed | Audience* |
| --- | --- | --- | --- | --- | --- |
| Websites<br>n = 53 | With references = 8<br>With links to other organizations = 2<br>Without external references/links = 43 | Private (individual/company): n = 20 (Academy, 2018; Agers; Aspiro, 2019; Center, 2021; Centers, 2021; A. Centre, 2021; L. H. S. Centre, 2021; Clinic, 2020; Diagnostics, 2021b; F. Health, 2020; M. Health, 2020; T. M. Health, 2021; healthline, 2021; Help, 2021; Hospital, 2021; InnerHour; Manual, 2019; Psycom, 2021a; Sigmund, 2021; Solutions, 2017)<br>Private (foundation/charity): n = 12 (Centre, 2017; Foundation, 2020; V. B. Health, 2021; healthychildren.org, 2021; S. F. Institute, 2021; Minds, 2021; Ontario; Sinai, 2021; Society; Stem4, 2012; TeensHealth, 2014; Understood, 2021)<br>Federal government: n = 10 (Association, 2018; Headspace, 2021; Healthdirect, 2020; inform, 2020; Mind, 2021; NHS, 2020a; Prevention, 2020; Psychiatry; raisingchildren.net.au, 2021; service, 2020) | GAD: n = 11 (BC, 2013; Center, 2021; L. H. S. Centre, 2021; M. Health, 2020; T. M. Health, 2021; V. B. Health, 2021; Hospital, 2021; Manual, 2019; Mind, 2021; Sinai, 2021; Van Ameringen et al., 2015)<br>GAD + various: n = 1 (Ontario)<br>Anxiety disorders: n = 23 (Academy, 2018; Aspiro, 2019; Association, 2013, 2018; A. Centre, 2021; Centre, 2017; Clinic, 2020; Diagnostics, 2021b; Healthdirect, 2020; healthline, 2021; InnerHour; S. F. Institute, 2021; KidsHealth, 2021; NHS, 2020a; Ontario, 2021; Psychiatry; raisingchildren.net.au, 2021; service, 2020; Society; Solutions, 2017; TeensHealth, 2014; Understood, 2021)<br>Anxiety: n = 17 (Agers; BC; Centers, 2021; Centre; Foundation, 2020; Headspace, 2021; F. Health, 2020; healthychildren.org, 2021; Help, 2021; | Yes: n = 47 (Academy, 2018; Aspiro, 2019; Association, 2013, 2018; BC; BC, 2013; Center, 2021; Centers, 2021; A. Centre, 2021; Centre, 2017; L. H. S. Centre, 2021; Centre; Clinic, 2020; Diagnostics, 2021b; Headspace, 2021; F. Health, 2020; M. Health, 2020; T. M. Health, 2021; V. B. Health, 2021; Healthdirect, 2020; healthline, 2021; healthychildren.org, 2021; Help, 2021; HeretoHelp, 2019; Hospital, 2021; inform, 2020; S. F. Institute, 2021; Manitoba; Manual, 2019; Mind, 2021; Minds, 2021; Moir et al., 2015; NHS, 2020a; Ontario, 2021; Prevention, 2020; Psychiatry; Psycom, 2021a; raisingchildren.net.au, 2021; service, 2020; Sigmund, 2021; Sinai, 2021; Society; Stem4, 2012; TeensHealth, 2014; Understood, 2021; Van Ameringen et al., 2015)<br>No: n = 6 (Agers; Foundation, 2020; InnerHour; KidsHealth, | Parents: n = 8 (Academy, 2018; Agers; Centers, 2021; Centre; Clark et al., 2018a; raisingchildren.net.au, 2021; service, 2020; Sigmund, 2021; Sinai, 2021)<br>Youth/children: n = 38 (Academy, 2018; Agers; Aspiro, 2019; Association, 2018; BC; BC, 2013; Center, 2021; Centers, 2021; Centre, 2017; L. H. S. Centre, 2021; Centre; Headspace, 2021; T. M. Health, 2021; V. B. Health, 2021; Healthdirect, 2020; healthychildren.org, 2021; HeretoHelp, 2019; Hospital, 2021; inform, 2020; S. F. Institute, 2021; KidsHealth, 2021; Manitoba; Manual, 2019; Mind, 2021; Minds, 2021; NHS, 2020a; Ontario, 2021; Prevention, 2020; Psychiatry; Psycom, 2021a; raisingchildren.net.au, 2021; service, 2020; |
|  |  | Provincial government: n = 9 (Association, 2013; BC; BC, 2013; HeretoHelp, 2019; KidsHealth, 2021; Manitoba; Ontario, 2021; Van Ameringen et al., 2015)<br>Academic: n = 2 (Centre; Moir et al., 2015) | HeretoHelp, 2019; inform, 2020; Manitoba; Minds, 2021; Moir et al., 2015; Psycom, 2021a; Sigmund, 2021; Stem4, 2012)<br>Anxiety and depression: n = 1 (Prevention, 2020) | 2021; Ontario; Solutions, 2017) | Sinai, 2021; Society; Stem4, 2012; TeensHealth, 2014; Understood, 2021)<br>NA: n = 14 (Association, 2013; A. Centre, 2021; Clinic, 2020; Diagnostics, 2021b; Foundation, 2020; F. Health, 2020; M. Health, 2020; healthline, 2021; Help, 2021; InnerHour; Moir et al., 2015; Ontario; Solutions, 2017; Van Ameringen et al., 2015) |
| Scientific studies<br>n = 5 | Cross-sectional = 3<br>Prospective = 1<br>Qualitative = 1 | Academic: n = 5 (Arcaro et al., 2017; Barzilay et al., 2020; Farmer et al., 2018; Narmandakh et al., 2020; Spence et al., 2018) | GAD: n = 2 (Barzilay et al., 2020; Spence et al., 2018)<br>Anxiety disorders: n = 2 (Farmer et al., 2018; Narmandakh et al., 2020)<br>Anxiety and mood disorders: n = 1 (Arcaro et al., 2017) | No: n = 4 (Barzilay et al., 2020; Farmer et al., 2018; Narmandakh et al., 2020; Spence et al., 2018)<br>Yes: n = 1 (Arcaro et al., 2017) | Youth: n = 5 (Arcaro et al., 2017; Barzilay et al., 2020; Farmer et al., 2018; Narmandakh et al., 2020; Spence et al., 2018) |
| Review pieces<br>n = 4 | Narrative review = 3<br>Descriptive study = 1 | Academic: n = 4 (Bandelow & Michaelis, 2015; Kendall PC, 2016; Scrandis, 2019; Wehry et al., 2015) | Anxiety disorders: n = 4 (Bandelow & Michaelis, 2015; Kendall PC, 2016; Scrandis, 2019; Wehry et al., 2015) | Yes: n = 2 (Scrandis, 2019; Wehry et al., 2015)<br>No: n = 2 (Bandelow & Michaelis, 2015; Kendall PC, 2016) | Youth: n = 3 (Bandelow & Michaelis, 2015; Kendall PC, 2016; Scrandis, 2019)<br>NA: n = 1 (Bandelow & Michaelis, 2015) |

**Abbreviations:** GAD = Generalized Anxiety Disorder; NA = Not Available
<sup>†</sup>for details on individual studies see Supplementary Table 1
\*some studies cover multiple audiences

Thirteen Information Sources regarding signs and symptoms were written specifically about GAD. The majority of Information Sources (n=41, 66%) broadly addressed “anxiety disorders”, while others specifically stated they were written for generalized anxiety and other types of anxiety disorders or mood disorders. Most Information Sources described anxiety according to its signs and symptomology, provided treatment options, resources for help-seeking or referral programs and management of anxiety.

The age ranges were generally grouped into the categories of children/youth and youth/early adulthood. Thirty-six of the included Information Sources specifically stated they addressed *youth* anxiety (age range 11-29 years). A notable number were tailored for parents of youth with anxiety (n=11, 18%).

Seventy-five percent (n=46) of the included Information Sources did not include references for their description of anxiety signs and symptomology. Only 11 sources included references embedded within the body of the text and four listed references at the end of their narrative description. Of the few Information Sources that used embedded references, 82% (n=9) of them were from an academic source (Figure 2).

**Figure 2:**
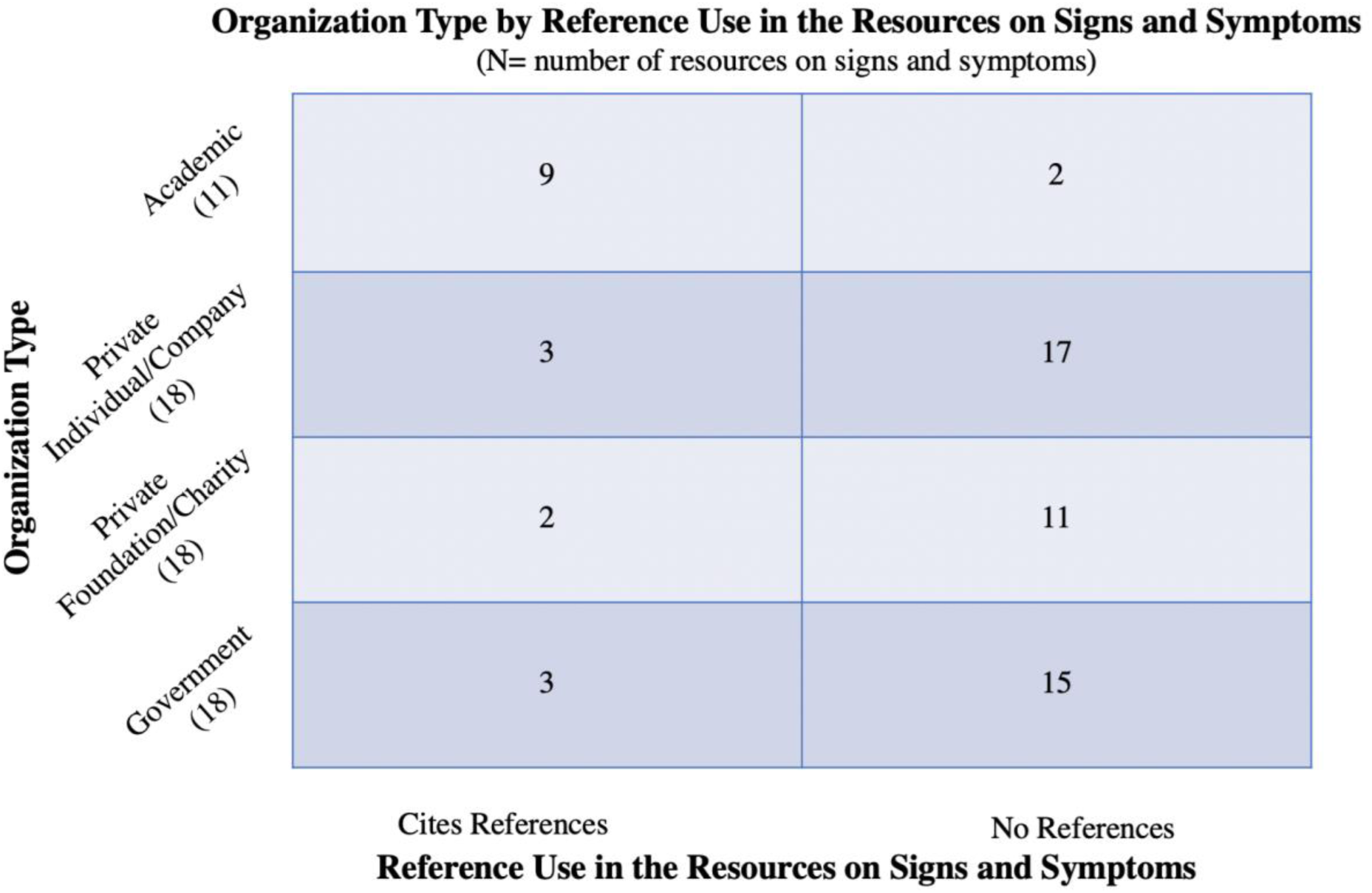

Only two (3%) Information Sources provided a link to an anxiety self-assessment resource, while 74% (n=46) provided information for additional resources including next steps for managing symptoms, links to external webpages, phone numbers to access therapists and crisis lines, promoted help-seeking, and 26% (n=16) did not provide any information for additional support.

A small number of the Information Sources (n=7, 11%) identified as businesses looking to attract clients by providing information on the signs and symptoms of anxiety, followed by a phone number or link to contact one of their paid therapists.

### What anxiety self-assessment resources are available online to youth?

#### Summary of resources available

Seventeen self-assessment resources were included from five countries (Table 2). However, the majority of resources came from the USA (n=9, 53%); with a notable number from Canada (n=4, 23%), Australia (n=2, 12%), India (n=1, 6%), and the UK (n=1, 6%). Most self-assessment resources were provided by private institutions (n=12, 71%), whether that be a company/individual or foundation/charity. Other organizations that provided self-assessment resources were governmental (n=4, 23%) and academic (n=1, 6 %).

**Table 2:** Summary of Study Characteristics for Self-Assessment Resources (Question #2)†

| Information Source | Types of Resources Provided* | Organization Type | Foundational Tool | Provides an Anxiety Score | Provides Self-Management Support | Specifically for Youth |
| --- | --- | --- | --- | --- | --- | --- |
| Websites<br>n = 13 | Links: n = 5<br>Contact information: n = 6<br>None: n = 4<br>Contact for internal psychologist: n = 3<br>External self-management support: n = 2 | Private (individual/company): n = 6 (Brisbane, 2017; Diagnostics, 2021a; Information, 2018; PsychCentral; PSYCOM, 2021b; Today, 2021)<br>Private (foundation/charity): n = 2 (B. D. Institute, 2021; Phone)<br>Federal government: n = 2 (America; NHS, 2020b)<br>Provincial government: n = 2 (BC; HereToHelp)<br>Academic: n = 1 (Pittsburgh, 2021) | GAD-7: n = 3 (Brisbane, 2017; Diagnostics, 2021a; Phone)<br>GAD-IV: n = 1 (America)<br>DSM-5: n = 1 (PsychCentral)<br>SCARED: n = 1 (Pittsburgh, 2021)<br>None: n = 7 (BC; HereToHelp; Information, 2018; B. D. Institute, 2021; NHS, 2020b; PSYCOM, 2021b; Today, 2021) | Yes: n = 8 (Brisbane, 2017; Diagnostics, 2021a; HereToHelp; Information, 2018; NHS, 2020b; Pittsburgh, 2021; PsychCentral; Today, 2021)<br>No: n = 5 (America; BC; B. D. Institute, 2021; Phone; PSYCOM, 2021b) | No: n = 13 (America; BC; Brisbane, 2017; Diagnostics, 2021a; HereToHelp; Information, 2018; B. D. Institute, 2021; NHS, 2020b; Phone; Pittsburgh, 2021; PsychCentral; PSYCOM, 2021b; Today, 2021) | Yes: n = 3 (BC; Phone; Pittsburgh, 2021)<br>No: n = 10 (America; Brisbane, 2017; Diagnostics, 2021a; HereToHelp; Information, 2018; B. D. Institute, 2021; NHS, 2020b; PsychCentral; PSYCOM, 2021b; Today, 2021) |
| Applications<br>n = 4 | Links: n = 3<br>Contact | Private (individual/company): n = 4 (InnerHour; LLC, | GAD-7: n = 3 (LLC, 2020, | Yes: n = 4 (InnerHour; LLC, 2020, | Yes: n = 2 (InnerHour; LLC, December 2016) | No: n = 4 (InnerHour; LLC, 2020, |
|  | information: n = 1<br>None: n = 1 | 2020, December 2016;<br>Taylor, 2015) | December 2016;<br>Taylor, 2015)<br>None: n = 1<br>(InnerHour) | December<br>2016; Taylor,<br>2015) | No: n = 2 (LLC,<br>2020; Taylor,<br>2015) | December<br>2016; Taylor,<br>2015) |

Abbreviations: GAD = Generalized Anxiety Disorder; NA = Not Available
†for details on individual studies see Supplementary Table 2
\*some provide multiple resources

The modality of self-assessments was either through a stand-alone website available for a desktop computer (n=13, 76%) or an App downloadable from an App Store (i.e., Google Play, iTunes) to a mobile device (n=4, 24%). All resources were available for use by youth, however only three of the 18 available resources explicitly stated being tailored for youth.

Every self-assessment resource included a questionnaire section in which the user would provide answers to a series of questions on a Likert scale. Six self-assessment resources (35%) explicitly stated that they used a previously validated anxiety assessment tool as a foundation. The foundational tools/manuals reported were the GAD-7 (Spitzer et al., 2006), GAD-IV (Newman et al., 2002), SCARED (Birmaher et al., 1999), and DSM-V (Substance & Mental Health Services, 2016). Nine used a validated tool as the foundation for their self-assessment questionnaire. Figure 3 shows a breakdown of the self-assessment resources (i.e., academic, governmental, or private) and their foundational tools.

**Figure 3:**
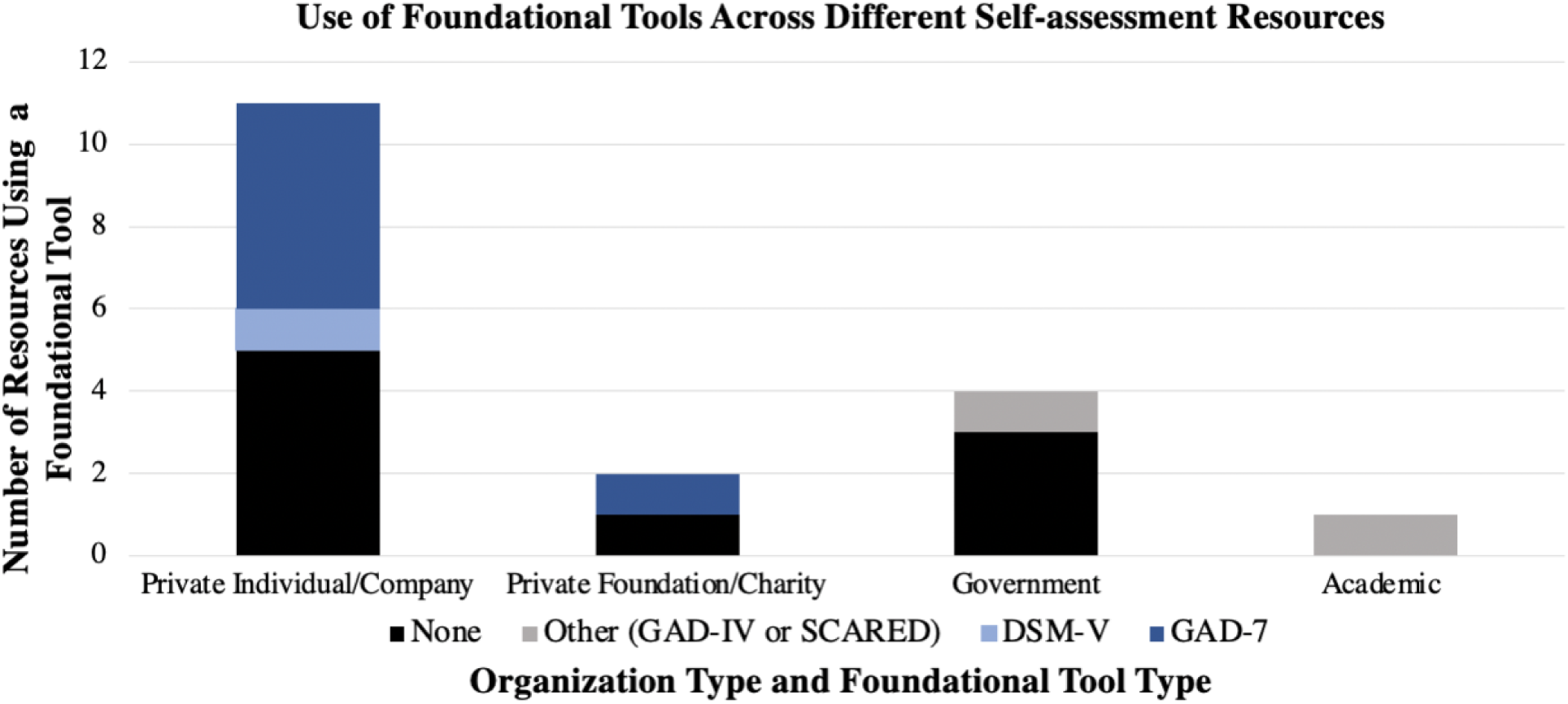

#### Post-assessment outputs

The majority of self-assessment resources provided a post-assessment number indicative of the user’s anxiety level (n=12, 67%). This number changed based on the answers that users provided to a pre-requisite questionnaire. Fifteen (83%) self-assessment resources provided a post-assessment statement specific to the user’s anxiety level. Six of these provided more than one sentence of descriptive text on the user’s anxiety level, with 17% (n=3) of the resources not changing the output based on the users’ responses (i.e., resources provided identical outputs when users would respond to all questions with high anxiety answers or low anxiety answers).

#### Post-assessment support provided

After completing the self-assessment, many of the resources (n=13, 72%) provided a wide range of supportive messages to the user. All self-assessment resources from a governmental institution (n=4, 22%) provided post-assessment support. Proportionally, self-assessment resources from private (n=13, 72%) and academic (n=1, 6%) institutions provided fewer post-assessment supports. Nearly every governmental self-assessment resource promoted help-seeking, provided links and contact information (n=3, 75%) (Figure 4). Comparatively, only half of private self-assessment resources promoted help-seeking and provided links post-assessment (n=7, 54%).

**Figure 4:**
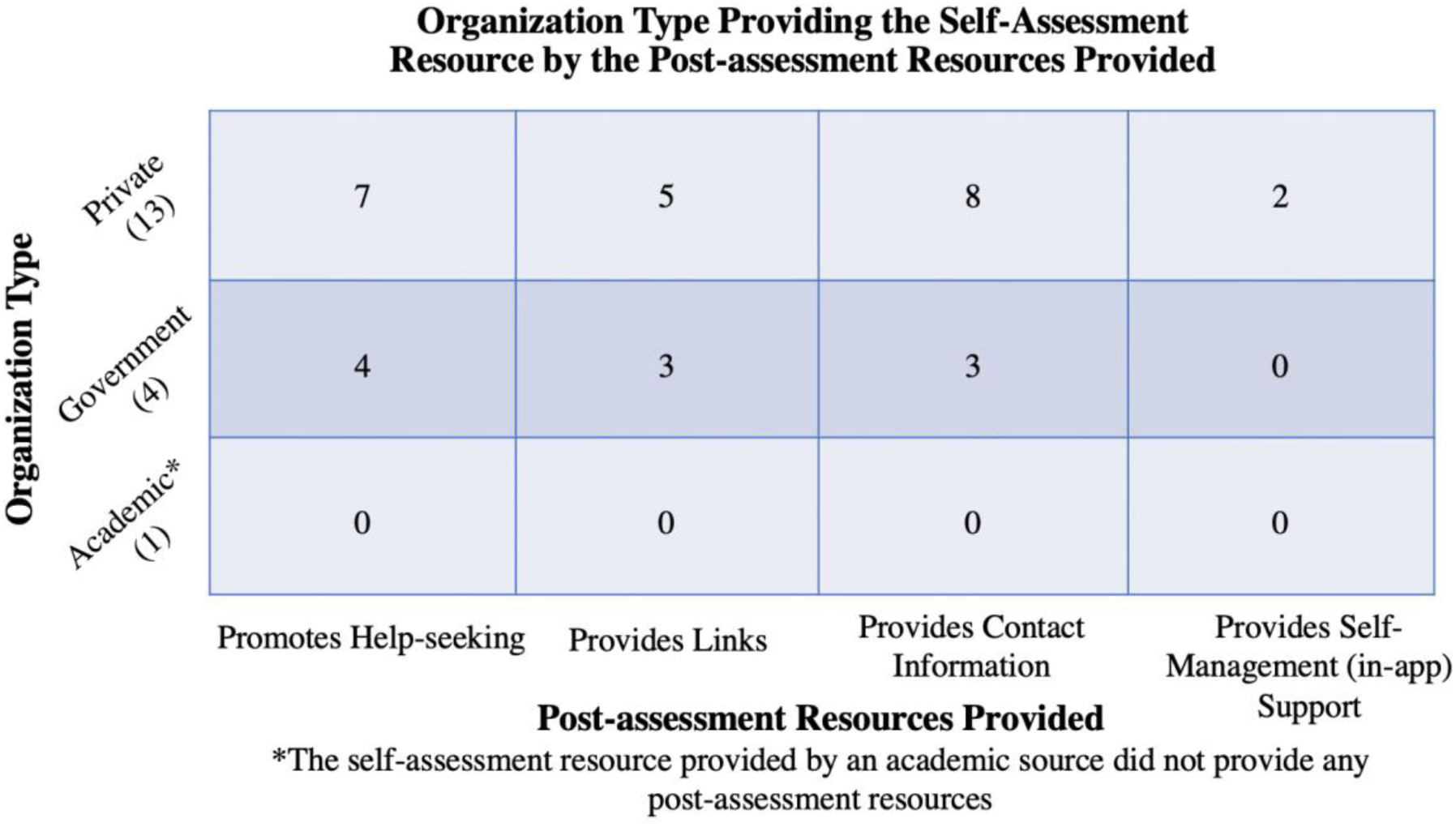

Promoting help-seeking, providing resources, and offering access to self-management support were common messages, with 72% (n=13) of the self-assessment resources providing at least one of those outputs. A reasonable number provided links and contact information such as an email, or phone number for a distress line or psychologist (n=8, 44% and n=11, 61%, respectively).

### What are youth’s experiences and information needs around seeking help for anxiety?

A total of 19 Information Sources relating to youth’s experiences and information needs around seeking help for anxiety were identified, 17 (Arcaro et al., 2017; Calear et al., 2020; Clark et al., 2018a, 2018b; Clark et al., 2020; Coles et al., 2015; Dayan, 2019; Elshire-Dulle, 2019; Grist et al., 2018; Haavik et al., 2019; Havinga et al., 2018; Langley et al., 2018; Leech et al., 2019; Moir et al., 2015; Purcell et al., 2015; Romanson, 2018; Summerhurst et al., 2017; Van Ameringen et al., 2015) through the primary literature search, one via Google Scholar (Gandhi et al., 2016), and one through the Internet search (Canada, 2018). Characteristics for each of these Information Sources are presented in Table 3. Participants ranged in age from 10 to 29 years. Information from participants was gathered through validated questionnaires (n=4, 21%), custom questionnaires (n=5, 26%), interviews (n=2, 11%) and a combination of these methods (n=8, 42%). The populations varied from medical/university students (n=5, 26%) and secondary students (n=3, 16%) to those diagnosed with anxiety disorders (n=4, 21%).

#### Barriers and predictors to seeking help for anxiety

With regards to help-seeking, several categories emerged relating to their experiences seeking in-person professional help, online help, and help in general (e.g., stigma associated with seeking help for anxiety disorders). Seeking professional help had mixed results from participants; in two Information Sources youth stated that they were unlikely to seek in person professional help if they were experiencing symptoms (Calear et al., 2020; Moir et al., 2015). Similarly, in two other Information Sources participants did not find talking (Summerhurst et al., 2017) or consulting with professionals to be helpful (Leech et al., 2019). Specifically, only 16% (n=3) of youth in one study with recent onset of mood/anxiety concerns found talking was the most helpful for recovery (Summerhurst et al., 2017).

Several Information Sources explored the use of the internet and social media by youth seeking help for anxiety disorders. A survey of youth in Australia rated information found on social media as unhelpful (38%) or neutral (33%) (Leech et al., 2019). Youth surveyed in the United Kingdom identified several advantages of using mental health Apps including being anonymous (65%), private (65%), and always being available (57%) (Grist et al., 2018). Although stigma was commonly referred to as a barrier, youth also described stigma relating to their overall experiences with help-seeking (Clark et al., 2020; Elshire-Dulle, 2019).

The most frequently reported barriers for help-seeking included accessibility to services (i.e., cost, living in smaller community, transition of care, transportation) and quality or perception of professional care. The cost and time to access services was often identified as a barrier (Elshire-Dulle, 2019; Haavik et al., 2019; Langley et al., 2018; Leech et al., 2019; Van Ameringen et al., 2015). A series of videos with young adults identified difficulties in transitioning from child-centered to adult services (Canada, 2018). This was often due to a lack of adequate transition from school-based to adult services, or in smaller communities’, adult services were not provided locally.

Low satisfaction and general lack in quality of care was also identified as a barrier for youth accessing mental health services for anxiety (Leech et al., 2019; Van Ameringen et al., 2015). Some youth were not convinced their anxiety symptoms were severe enough and therefore would not benefit from professional help or that unnecessary medication would be prescribed (Langley et al., 2018; Van Ameringen et al., 2015).

Predictors of service use for anxiety centered around two main themes: gender and awareness of their symptoms or anxiety scores. Across several Information Sources that assessed youth anxiety, those with higher scores were associated with greater intentions to consult professional help (Calear et al., 2020; Grist et al., 2018; Leech et al., 2019; Moir et al., 2015; Romanson, 2018; Van Ameringen et al., 2015). All of these Information Sources with the exception of one used a validated scale to assess anxiety (Calear et al., 2020; Grist et al., 2018; Leech et al., 2019; Moir et al., 2015; Romanson, 2018). The other study used a customized online survey (Van Ameringen et al., 2015). Four Information Sources found that females were much more likely than males to access professional services, including social media resources and university services (Arcaro et al., 2017; Calear et al., 2020; Haavik et al., 2019; Havinga et al., 2018).

#### Information needs

Only five Information Sources regarding information needs were identified. When information needs were examined the context was often with regards to seeking help from online sources (Leech et al., 2019; Van Ameringen et al., 2015). Two information sources (Clark et al., 2018b; Dayan, 2019) discussed needing information in schools being accessible and visible to youth in an effort to narrow the gap of unmet mental health needs (Canada, 2018).

## Discussion

As youth are highly vulnerable to sustained stressors, it is not surprising that the COVID-19 pandemic has been associated with a rise in the prevalence of youth anxiety disorders and an increase in accessible mental health supports (Courtney et al., 2020). Moreover, in Canada adolescents experience higher rates of accessing health services for mood and anxiety disorders than any other age group (McRae et al., 2016b). By reviewing the existing literature and available online sources, this systematic environmental scan provides a comprehensive overview of 1) information available online to Canadian youth about the signs and symptoms of anxiety, 2) resources available online to self-assess anxiety and 3) youth’s information needs and experiences help-seeking for anxiety. Specifically, 99 Information Sources were included from the Internet and primary literature and contextualized through discussions with youth collaborator groups.

In addressing the three questions of this systematic environmental scan it was highlighted that: (1) there are not many online anxiety resources developed specifically for Canadian youth, and of the limited information for youth, little is developed with youth perspectives in mind; (2) much of the information that is available online, where most youth search, is not clearly backed by academic references; and (3) there is a lack of research on the information needs of youth seeking help for anxiety. Below, we highlight implications for improvement of online anxiety information sources for youth. Results from this study, and new lines of research in this area, will be valuable in informing those producing information in this domain.

### Improving the Youth-friendliness of Resources

Very few Information Sources addressing the signs and symptoms of anxiety and self-assessment resources were deemed ‘youth-friendly’. Youth collaborators suggested that youth desire straight-forward, accessible tools that ensure the privacy of their identifying information. Further, straight-forward descriptions in lay language are preferable. Some self-assessment resources required youth to provide personal information before accessing support options. The privacy concerns raised by some youth collaborators mirror the barriers to seeking mental health information and support online which include anonymity and confidentiality (Pretorius et al., 2019). Further, a minority of self-assessment resources appear to be designed with the intention of attracting new clients to for-profit services. For low-income youths, tools designed in this way limit access to anxiety resources and support. The literature and discussions with youth collaborators emphasize the provision of accessible supports as a priority of the online mental health resources and tools youth access (Kenny et al., 2016). Tools providing paid post-assessment support should consider including links to free and evidence-based anxiety information webpages and emergency supports. Resources for self-assessment and the signs and symptoms of anxiety varied markedly in their provision of additional information post-assessment. Qualitative differences in the accessibility of post-assessment support existed in the tools provided by governmental and private sources: each self-assessment resource from a governmental source provided more forms of post-assessment support (i.e., promoted help-seeking, provided links, or contact information) than those from private sources. Further, youth collaborators highlighted a need for the inclusion of location-specific online supports outlined within these resources to facilitate next steps in seeking support. Youth requested that resources include information on starting conversations about their self-assessment results with others (i.e., parents, peers, and healthcare providers). Attention to the delivery of accessible, youth-friendly, and location-specific information and support should be a requirement of all resources.

### Making Credible and Evidence-based Information a Priority

While the credibility of identified online resources was occasionally substantiated by evidence-based references or a validated foundational tools (i.e., GAD-7), this was not always the case. Less than half of identified self-assessment resources rely on a validated foundational tool, thus drawing into question the credibility and trustworthiness of the outcomes presented to users. Moreover, a handful of resources ‘assessing anxiety levels’ did not change their output depending on the users’ responses to the assessment questions. It seems unlikely that these outputs are evidence-based. This finding is consistent with themes developed from a qualitative study on mental health information-searching: several participants described searches for online mental health information as yielding “superficial” and “vague or inconclusive” information and had concerns about the sources of such information (Lal et al., 2018). Those developing self-assessment resources should reference previously validated tools as a foundation for their construction and provide outputs specific to the users’ responses. Discussion with youth collaborators highlighted the cues youth may rely on when assessing the trustworthiness of a resource, such as reported alignment with a credible organization, a professional user interface, and the medium of the information (i.e., webpage, social media). These findings mirror the literature, which outlines that youth trust online health information from websites more than social media sites and rely on heuristics such as institutional logos to assess trustworthiness (Freeman et al., 2020). Unfortunately, these cues are not always indicative of evidence-based information and were often not evident in the resources identified. Further, only 25% of the resources describing the signs and symptoms of anxiety included academic references. The disparity in the use and reporting of validated foundational tools and references needs to be addressed, as youth desire credible and accurate online mental health information (Bradley et al., 2012; Kenny et al., 2016).

### Information Needs of Youth

To develop effective and credible tools for the assessment and management of youth anxiety we need to know what youth seek when it comes to information about their anxiety. Through our search of the primary literature and the Internet there was a paucity of Information Sources on this topic. When emerging adults were asked about information needs they identified several resources that would ease transition from school-based to community-based services (Canada, 2018). Moreover, youth collaborators provided valuable insights about the type and format of information they are seeking. These earlier interviews and discussions with our youth collaborators demonstrate that, if asked, youth have preferences and opinions about information that they desire (Canada, 2018). However, this is not reflected in assessment and management resources available to youth about their anxiety needs. Given the effect of the COVID-19 pandemic on youth mental health, responding to this need in a timely manner has become crucial (Courtney et al., 2020).

### Youth Engagement in the Development of Future Resources

The importance of patient-oriented research has been recognized as an integral component to capture the lived experiences of those experiencing specific conditions and interactions with the healthcare system (Manafo et al., 2018; Research, 2012). Patient-oriented research anticipates that partnering patients with researchers will enhance research quality and will “improve healthcare policies and practices across the system, ultimately improving health outcomes” (Research, 2019). Patient-oriented research emphasizing youth voices may reveal valuable insights in the development of youth-friendly resources with the potential for greater relevance and impact (J.C.F.S. Health, 2021). Therefore, it is imperative that youth are included in the development of future resources for their anxiety needs and are consulted on their help-seeking preferences.

#### Strengths and Limitations

By including youth collaborators in the co-creation of the research question and discussion of findings, we expect that the results of the work will guide future efforts that yield outcomes of greater relevance to youth. Additionally, our systematic approach to screening and verification of identified Information Sources ensured a rigorous collation of available Information Sources.

While we set out to provide a comprehensive overview of available Information Sources regarding the signs and symptoms of anxiety, self-assessment resources, help-seeking preferences, and information needs, we may not have captured it all. Although a systematic process was employed, websites may have been missed due to the limited number of key terms, Google indexing and search location preferences. Additionally, due to the dynamic nature of the internet, relevant websites may have been made accessible after our primary searches were completed. As we tried to mimic the online search behaviors of *Canadian* youth, the available websites are limited to location caching within Google (i.e., running the same search in the United Kingdom would elicit different webpage results) and thus the generalizability of the results is limited for those outside of Canada. Further, restricting our search to the Google search engine may have been a limitation as novel resources could be accessible through other popular search engines (e.g., Bing, Yahoo), however, Google is the most popular, most widely used search engine (Johnson, 2021).

Notably, the focus of this study was GAD, restricting the generalizability of findings to other anxiety and mood disorders.

#### Future work

Identifying the gaps in the available resources will help guide primary research as well as knowledge translation efforts to support youth in accessing the information they need for recognizing and subsequently managing anxiety. Key considerations and messages from youth collaborators contextualizing these results, provides informed guidance for the continued development of youth anxiety self-assessment resources.

## Conclusion

Currently, the format of online information available to youth about anxiety disorders is primarily website-based. Self-assessment resources vary markedly in the support they provide and their ability to meet youth preferences. Limited information exists around the information needs and anxiety help-seeking preferences of youth. As the size of the body of information in this domain continues to increase, coupled with the impact the current pandemic is having on youth’s mental health, the need to periodically monitor and assess available information is crucial. Collectively, the findings of this study highlight the need for the development of online anxiety resources informed by youth perspectives and preferences. Researchers and healthcare providers engaging youth in the development process can support the creation of resources of greater relevance and usefulness to youth, increasing their accessibility to care.

## Data Availability

All data produced in the present work are contained in the manuscript and supplementary files

## Acknowledgements

The authors would like to extend their sincerest thanks to the youth collaborators (the Alberta Children’s Hospital Child and Youth Advisory Council, the Stollery Children’s Hospital Youth Advisory Council, and the National KidsCan Young Person’s Advisory Group) who supported the project and provided insightful discussions and comments on the results of the environmental scan.

## Appendix 1: Electronic Search Strategie

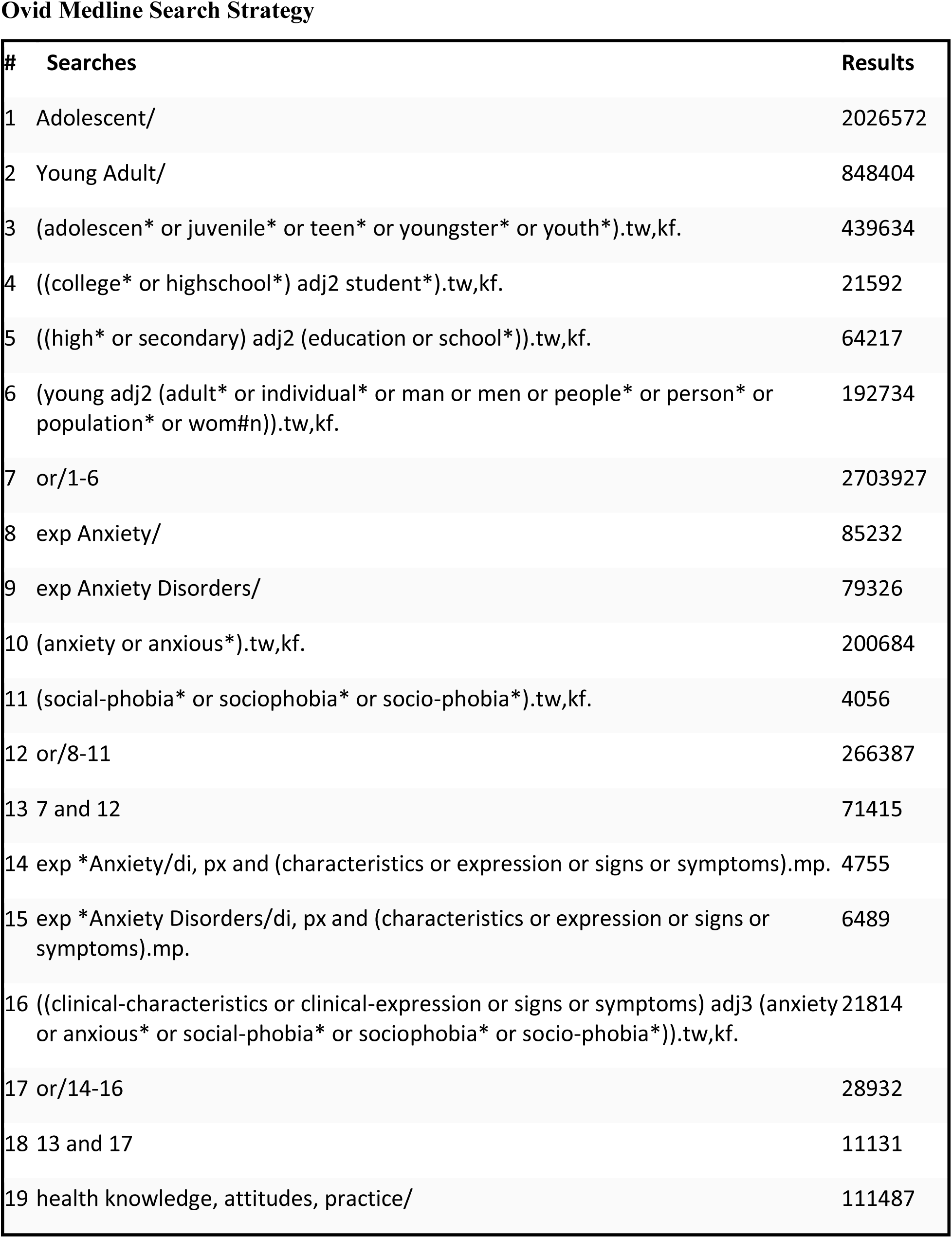
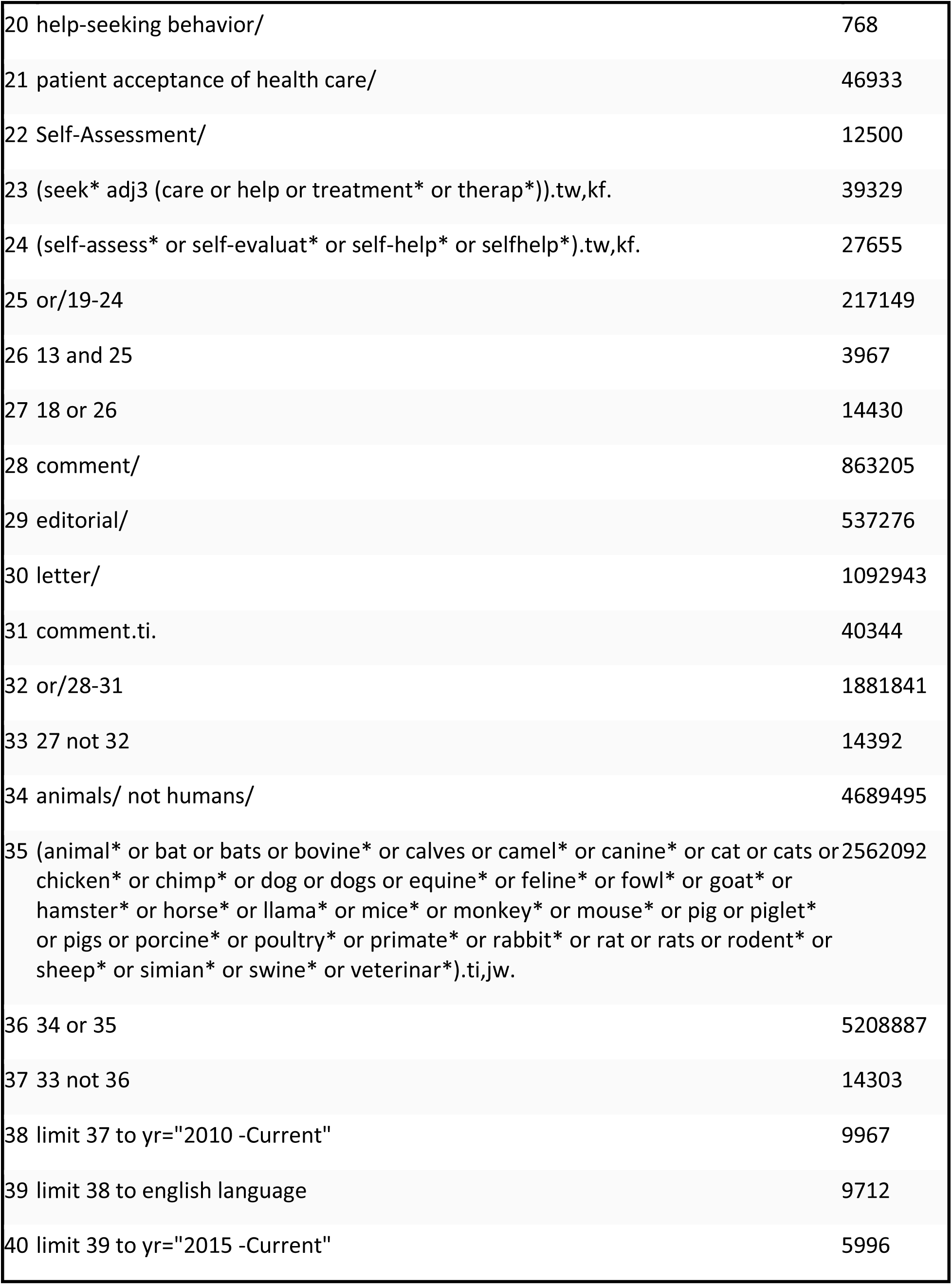
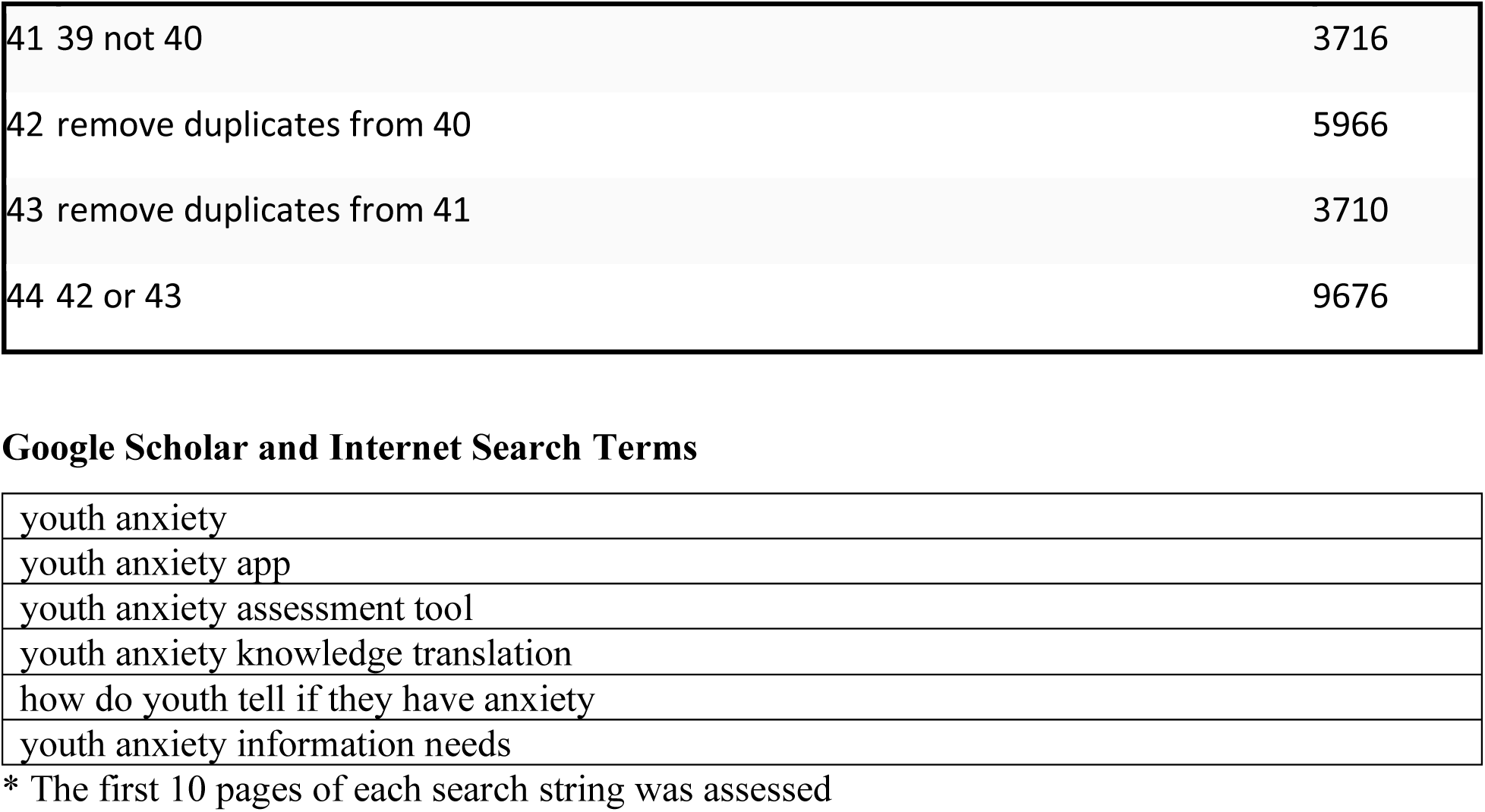

Ovid Medline Search Strategy
| # | Searches | Results |
| --- | --- | --- |
| 1 | Adolescent/ | 2026572 |
| 2 | Young Adult/ | 848404 |
| 3 | (adolescen* or juvenile* or teen* or youngster* or youth*).tw,kf. | 439634 |
| 4 | ((college* or highschool*) adj2 student*).tw,kf. | 21592 |
| 5 | ((high* or secondary) adj2 (education or school*)).tw,kf. | 64217 |
| 6 | (young adj2 (adult* or individual* or man or men or people* or person* or population* or wom#n)).tw,kf. | 192734 |
| 7 | or/1-6 | 2703927 |
| 8 | exp Anxiety/ | 85232 |
| 9 | exp Anxiety Disorders/ | 79326 |
| 10 | (anxiety or anxious*).tw,kf. | 200684 |
| 11 | (social-phobia* or sociophobia* or socio-phobia*).tw,kf. | 4056 |
| 12 | or/8-11 | 266387 |
| 13 | 7 and 12 | 71415 |
| 14 | exp *Anxiety/di, px and (characteristics or expression or signs or symptoms).mp. | 4755 |
| 15 | exp *Anxiety Disorders/di, px and (characteristics or expression or signs or symptoms).mp. | 6489 |
| 16 | ((clinical-characteristics or clinical-expression or signs or symptoms) adj3 (anxiety or anxious* or social-phobia* or sociophobia* or socio-phobia*)).tw,kf. | 21814 |
| 17 | or/14-16 | 28932 |
| 18 | 13 and 17 | 11131 |
| 19 | health knowledge, attitudes, practice/ | 111487 |

|  |  |
| --- | --- |
| 20 help-seeking behavior/ | 768 |
| 21 patient acceptance of health care/ | 46933 |
| 22 Self-Assessment/ | 12500 |
| 23 (seek* adj3 (care or help or treatment* or therap*)).tw,kf. | 39329 |
| 24 (self-assess* or self-evaluat* or self-help* or selfhelp*).tw,kf. | 27655 |
| 25 or/19-24 | 217149 |
| 26 13 and 25 | 3967 |
| 27 18 or 26 | 14430 |
| 28 comment/ | 863205 |
| 29 editorial/ | 537276 |
| 30 letter/ | 1092943 |
| 31 comment.ti. | 40344 |
| 32 or/28-31 | 1881841 |
| 33 27 not 32 | 14392 |
| 34 animals/ not humans/ | 4689495 |
| 35 (animal* or bat or bats or bovine* or calves or camel* or canine* or cat or cats or chicken* or chimp* or dog or dogs or equine* or feline* or fowl* or goat* or hamster* or horse* or llama* or mice* or monkey* or mouse* or pig or piglet* or pigs or porcine* or poultry* or primate* or rabbit* or rat or rats or rodent* or sheep* or simian* or swine* or veterinar*).ti,jw. | 2562092 |
| 36 34 or 35 | 5208887 |
| 37 33 not 36 | 14303 |
| 38 limit 37 to yr="2010 -Current" | 9967 |
| 39 limit 38 to english language | 9712 |
| 40 limit 39 to yr="2015 -Current" | 5996 |
| 41 39 not 40 | 3716 |
| 42 remove duplicates from 40 | 5966 |
| 43 remove duplicates from 41 | 3710 |
| 44 42 or 43 | 9676 |

Google Scholar and Internet Search Terms
|  |
| --- |
| youth anxiety |
| youth anxiety app |
| youth anxiety assessment tool |
| youth anxiety knowledge translation |
| how do youth tell if they have anxiety |
| youth anxiety information needs |

\* The first 10 pages of each search string was assessed

## Appendix 2: Eligibility Criteria

|  | **Include** | **Exclude** |
| --- | --- | --- |
| **Information Source Type** | Primary research (observational, quantitative, qualitative, and mixed methods), knowledge syntheses products (narrative, systematic, and overviews of reviews), and discussion papers/documents.<br>Grey literature including Canadian websites and online sources describing self-assessment tool and apps for youth anxiety. | Editorials, letters, and opinion pieces, abstracts only.<br>All intervention studies. |
| **Language** | English | Languages other than English |
| **Country** | Developed countries<sup>(Prospects, 2014)</sup>. | Website, handouts, educational material from non-governmental, non-registered practitioner, non-health and non-tertiary institutions. |
| **Publication date** | 2015 onwards. | Pre-2015. |
| **Population** | Youth and children between the ages of 15-24 years with no previous triggering event to cause anxiety<sup>((UNDESA), 2008)</sup>. | Persons older than 25 years of age.<br>Persons with chronic conditions such as epilepsy, autism, attention deficit disorder, chronic pain, cancer, concussion, etc.<br>Unique populations such as refugees, youth brides, peer victimization /bullying, and institutional deprivation. |
| **Exposure** | Anxiety | NA |
| **Comparator** | NA | NA |
| **Outcomes** | Signs and symptoms of youth experiencing general anxiety disorder, or<br>Techniques or methods/tools used to recognize/self-assess anxiety and when to seek help, or | Studies that do not report on the signs and symptoms of general anxiety disorder.<br>Studies reporting on anxiety as a result of managing chronic illness, substance abuse, previous suicide attempts, bullying/victimization, divorce, trauma, : racial discrimination, food/home insecurity during childhood or |
|  | Youth experiences and information needs around seeking help for general anxiety disorder.<br>- Self-reports of the type including quantity, timing, and delivery mode of information desired by youth. | other anxiety related disorders (e.g., obsessive-compulsive, panic, or post-traumatic stress disorders, social phobia, social anxiety disorder, needle/pain anxiety, etc.). |

## Appendix 3: Definitions

| Term | Definition |
| --- | --- |
| Information Source | Collective term referring to primary literature, Google, and Internet sources |
| App | An application downloadable from an App Store (i.e., Google Play, iTunes) to a mobile device |
| Website | The assessment is completed through a stand-alone website available for a desktop computer |
| Foundational Tools of the Self-Assessment Resources | Yes, has a foundational tool: The self-screening resource either explicitly states GAD-7 or the DSM-V has been used as the foundational tool to create this resource or the tool includes all questions from the GAD-7 or all diagnostic questions from the DSM-V but does not report it as the foundational tool<br>No, does not have a foundational tool: The self-screening resource does not meet the requirements to qualify for the “yes, has a foundational tool definition” |
| Avenues for Next Steps | Promotes help-seeking: Post-assessment, the resource explicitly suggests that the user seek help from their family, physician, or other anxiety self-management aids (e.g. "It seems like you have anxiety, you should reach out to your healthcare provider")<br>Provides resources: Post-assessment, the resource provides the user with links to anxiety information sheets, the contact information for a distress line or psychologist, or other forms of support.<br>Provides self-management (in-app) support: Post-assessment, the resource offers support to users within the application or webpage of the company that provided the assessment. This must include the option for the user to self-manage anxiety through the application or webpage. This can also include contact (through the application or webpage) with healthcare professionals associated with the organization that administered the self-screening.<br>Provides links: The self-screening resource provides links to anxiety information sheets or webpages on signs, symptoms, and other facets of anxiety<br>Provides contact information: The self-screening resource provides contact information such as a cell phone number or email (distress line or psychologist's contact information)<br>Provides external self-management support: Provides links to third-party applications that assist with anxiety self-management |
|  | Definitions for the Types of Resources Provided by the Self-Screening Resource:<br>Links: Provides website links to anxiety information sheets or webpages on signs, symptoms, and other facets of anxiety<br>Contact Information: Provides contact information, such as a cell phone number or email for a distress line or psychologist<br>Definitions for the Types of Outputs of the Self-Screening Resources:<br>Number: The resource provides a number signifying an anxiety score post-assessment<br>Statement: Source provides a one sentence statement that the user may have anxiety<br>Descriptive Text: Source provides descriptive text about the user's level of anxiety beyond a one sentence statement (see "statement" definition above) |

## References

(UNDESA), U. N. D. o. E. a. S. A. (2008). Definition of Youth. Retrieved April 21, 2021 from https://www.un.org/esa/socdev/documents/youth/fact-sheets/youth-definition.pdf

Academy, N. (2018). How to Recognize Anxiety in Teenagers. Retrieved April 2, 2021 from https://www.newportacademy.com/resources/mental-health/how-to-recognize-teen-anxiety-disorders/

Agers, S. Anxiety - What Every Young Person Should Know. Retrieved April 2, 2021 from https://www.screenagersmovie.com/tech-talk-tuesdays/anxiety-what-every-young-person-should-know

America, A. D. A. o. Screening for Generalized Anxiety Disorder (GAD). Retrieved April 4, 2021 from https://adaa.org/screening-generalized-anxiety-disorder-gad

Arcaro, J., Summerhurst, C., Vingilis, E., Wammes, M., & Osuch, E. (2017, 09). Presenting concerns of emerging adults seeking treatment at an early intervention outpatient mood and anxiety program. Psychology Health & Medicine, 22(8), 978–986. https://doi.org/https://dx.doi.org/10.1080/13548506.2016.1248449

Aspiro. (2019). 13 Signs of Anxiety in Young Adults and Teenagers. Retrieved March 27, 2021 from https://aspiroadventure.com/blog/13-signs-of-anxiety-in-teens-and-young-adults/

Association, C. M. H. (2013). *Anxiety Disorders*. Retrieved March 28, 2021 from https://cmha.bc.ca/documents/anxiety-disorders/

Association, C. M. H. (2018). *Children, Youth, and Anxiety*. Retrieved March 27, 2021 from https://cmha.ca/documents/children-youth-and-anxiety

Bandelow, B., & Michaelis, S. (2015, Sep). Epidemiology of anxiety disorders in the 21st century. Dialogues Clin Neurosci, 17(3), 327–335. https://doi.org/10.31887/DCNS.2015.17.3/bbandelow

Bandelow, B., Michaelis, S., & Wedekind, D. (2017, Jun). Treatment of anxiety disorders. Dialogues Clin Neurosci, 19(2), 93–107. https://doi.org/10.31887/DCNS.2017.19.2/bbandelow

Barzilay, R., White, L. K., Moore, T. M., Calkins, M. E., Taylor, J. H., Patrick, A., Huque, Z. M., Young, J. F., Ruparel, K., Pine, D. S., Gur, R. C., & Gur, R. E. (2020, Jun 05). Association of anxiety phenotypes with risk of depression and suicidal ideation in community youth. Depression & Anxiety, 05, 05. https://doi.org/https://dx.doi.org/10.1002/da.23060

BC, F. *Anxiety Self-check*. Retrieved April 4, 2021 from https://foundrybc.ca/quiz/anxiety-self-check/?return_page=1292

BC, T. C. I. a. S. P. C. o. (2013). Anxiety. Retrieved March 27, 2021 from https://youthinbc.com/youth-issues-2/mental-health/anxiety/

Birmaher, B., Brent, D. A., Chiappetta, L., Bridge, J., Monga, S., & Baugher, M. (1999, Oct). Psychometric properties of the Screen for Child Anxiety Related Emotional Disorders (SCARED): a replication study. Journal of the American Academy of Child & Adolescent Psychiatry, 38(10), 1230–1236. https://doi.org/10.1097/00004583-199910000-00011

Bradley, K. L., Robinson, L. M., & Brannen, C. L. (2012, 2012/02/01). Adolescent help-seeking for psychological distress, depression, and anxiety using an Internet program. International Journal of Mental Health Promotion, 14(1), 23–34. https://doi.org/10.1080/14623730.2012.665337

Brisbane, A. H. (2017). Self Assessment Quiz. Retrieved April 4, 2021 from https://anxietyhouse.com.au/appointments/self-assessment-quiz/

Calear, A. L., Batterham, P. J., Torok, M., & McCallum, S. (2020, Mar 16). Help-seeking attitudes and intentions for generalised anxiety disorder in adolescents: the role of anxiety literacy and stigma. European Child & Adolescent Psychiatry, 16, 16. https://doi.org/https://dx.doi.org/10.1007/s00787-020-01512-9

Canada, M. H. C. o. (2018). Emerging Adults Seek Change in Mental Health Services. Retrieved March 27, 2021 from https://www.youtube.com/playlist?list=PL2NuAPXp8ohbUt1WW0ga4afMYMmRSr7W Z

Canada, S. (2020). Impacts on Mental Health. Retrieved March 10, 2021 from https://www150.statcan.gc.ca/n1/pub/11-631-x/2020004/s3-eng.htm

Center, U. o. R. M. (2021). Generalized Anxiety Disorder (GAD) in Children and Teens. Retrieved April 2, 2021 from https://www.urmc.rochester.edu/encyclopedia/content.aspx?ContentTypeID=90&ContentID=P02565

Centers, E. T. (2021). Generalized Anxiety Disorder in Teens. Retrieved April 4, 2021 from https://evolvetreatment.com/parent-guides/anxiety/

Centre, A. (2021). Frequent Questions about Anxiety, Anxiety Disorder, Anxiety Attacks (Panic Attacks), etc. Retrieved March 27, 2021 from https://www.anxietycentre.com/anxiety-faq.shtml

Centre, F. (2017). Anxiety in Children and Youth. Retrieved March 27, 2021 from https://www.famcentre.ca/anxiety-in-children-and-youth/

Centre, L. H. S. (2021). Information About Mood & Anxiety. Retrieved March 27, 2021 from https://www.lhsc.on.ca/femap-first-episode-mood-and-anxiety-program/information-about-mood-anxiety

Centre, M. U. H. Anxious children: Treating a crippling disorder. Retrieved March 28, 2021 from https://www.thechildren.com/health-info/conditions-and-illnesses/anxious-children-treating-crippling-disorder

Charlton, P., Doucet, S., Azar, R., Nagel, D. A., Boulos, L., Luke, A., Mears, K., Kelly, K. J., & Montelpare, W. J. (2019). The use of the environmental scan in health services delivery research: a scoping review protocol. BMJ Open, 9(9), e029805. https://doi.org/10.1136/bmjopen-2019-029805

Clark, L. H., Hudson, J. L., Dunstan, D. A., & Clark, G. I. (2018a, Sep). Barriers and facilitating factors to help-seeking for symptoms of clinical anxiety in adolescent males [Empirical Study; Interview; Qualitative Study]. Australian Journal of Psychology, 70(3), 225–234. https://doi.org/http://dx.doi.org/10.1111/ajpy.12191

Clark, L. H., Hudson, J. L., Dunstan, D. A., & Clark, G. I. (2018b, Oct). Capturing the attitudes of adolescent males’ towards computerised mental health help-seeking [Empirical Study; Interview; Focus Group; Qualitative Study]. Australian Psychologist, 53(5), 416–426. https://doi.org/http://dx.doi.org/10.1111/ap.12341

Clark, L. H., Hudson, J. L., & Haider, T. (2020). Anxiety Specific Mental Health Stigma and Help-Seeking in Adolescent Males. Journal of Child & Family Studies, 29(7), 1970–1981. https://doi.org/10.1007/s10826-019-01686-0

Clinic, C. (2020). Anxiety Disorders. Retrieved April 2, 2021 from https://my.clevelandclinic.org/health/diseases/9536-anxiety-disorders

Coles, M. E., Coleman, S. L., & Schubert, J. (2015, Mar). College students’ recommendations for dealing with anxiety disorders [Empirical Study; Quantitative Study]. International Journal of Mental Health Promotion, 17(2), 68–77. https://doi.org/http://dx.doi.org/10.1080/14623730.2015.1005969

Courtney, D., Watson, P., Battaglia, M., Mulsant, B., & Szatmari, P. (2020). COVID-19 Impacts on Child and Youth Anxiety and Depression: Challenges and Opportunities. The Canadian Journal of Psychiatry, 65(10), 688–691. https://doi.org/10.1177/0706743720935646

Dayan, J. (2019). Attitude towards mental illness and help seeking behavior among college students:A pre-post design [Dissertation Empirical Study; Quantitative Study]. Dissertation Abstracts International: Section B: The Sciences and Engineering, 80(3-B(E)), No Pagination Specified. https://resolver.library.ualberta.ca/resolver?sid=OVID:psycdb&id=pmid:&id=doi:&issn=0419-4217&isbn=978-0438601611&volume=80&issue=3-B%28E%29&spage=No&pages=No+Pagination+Specified&date=2019&title=Dissertation+Abstracts+International%3A+Section+B%3A+The+Sciences+and+Engineering&atitle=Attitude+towards+mental+illness+and+help+seeking+behavior+among+college+students%3AA+pre-post+design.&aulast=Dayan&pid=%3Cauthor%3EDayan%2C+Jasmine%3C%2Fauthor%3E%3CAN%3E2018-65236-180%3C%2FAN%3E%3CDT%3EDissertation%3C%2FDT%3E

de Girolamo, G., Dagani, J., Purcell, R., Cocchi, A., & McGorry, P. D. (2012, Mar). Age of onset of mental disorders and use of mental health services: needs, opportunities and obstacles. Epidemiology & Psychiatric Science, 21(1), 47–57. https://doi.org/10.1017/s2045796011000746

Diagnostics, M. (2021a). Find Out If You Have Anxiety. Retrieved April 4, 2021 from https://www.mind-diagnostics.org/anxiety-test?utm_source=AdWords&utm_medium=Search_PPC_c&utm_term=anxiety%20disorder_b&utm_content=88328057960&network=g&placement=&target=&matchtype=b&utm_campaign=6453264419&ad_type=mind-diagnostics&adposition=&gclid=Cj0K

Diagnostics, M. (2021b). Find Out If You Have Anxiety. Retrieved March 27, 2021 from https://www.mind-diagnostics.org/anxiety-test?utm_source=AdWords&utm_medium=Search_PPC_c&utm_term=anxiety%20disorder_b&utm_content=88328057960&network=g&placement=&target=&matchtype=b&utm_campaign=6453264419&ad_type=mind-diagnostics&adposition=&gclid=Cj0K

Elliott, S. A. (2021, May 4). Establishing priorities in child health research: giving parents and youth a voice. Pediatric Academics Societies, Virtual.

Elshire-Dulle, J. (2019). The prevalence of and issues associated with the help seeking behavior among college student-athletes [Dissertation Empirical Study; Qualitative Study; Quantitative Study]. Dissertation Abstracts International Section A: Humanities and Social Sciences, 80(8-A(E)), No Pagination Specified. https://resolver.library.ualberta.ca/resolver?sid=OVID:psycdb&id=pmid:&id=doi:&issn=0419-4209&isbn=978-1392044339&volume=80&issue=8-A%28E%29&spage=No&pages=No+Pagination+Specified&date=2019&title=Dissertation+Abstracts+International+Section+A%3A+Humanities+and+Social+Sciences&atitle=The+prevalence+of+and+issues+associated+with+the+help+seeking+behavior+among+college+student-athletes.&aulast=Elshire-Dulle&pid=%3Cauthor%3EElshire-Dulle%2C+Jamie%3C%2Fauthor%3E%3CAN%3E2019-41138-294%3C%2FAN%3E%3CDT%3EDissertation%3C%2FDT%3E

Farmer, C., Thienemann, M., Leibold, C., Kamalani, G., Sauls, B., & Frankovich, J. (2018, 08 01). Psychometric Evaluation of the Caregiver Burden Inventory in Children and Adolescents With PANS [Research Support, N.I.H., Intramural

Research Support, Non-U.S. Gov’t]. Journal of Pediatric Psychology, 43(7), 749–757. https://doi.org/https://dx.doi.org/10.1093/jpepsy/jsy014

Foundation, B. a. B. R. (2020). Anxiety FAQs. Retrieved April 2, 2021 from https://www.bbrfoundation.org/faq/frequently-asked-questions-about-anxiety?gclid=CjwKCAiAnvj9BRA4EiwAuUMDf4uY-2P1fnBqaJJ29vn-VNgfeO3w7zV68OWpkgA0AXqZZjUGjGw-yhoCm1EQAvD_BwE

Freeman, J. L., Caldwell, P. H. Y., & Scott, K. M. (2020, Jun). The Role of Trust When Adolescents Search for and Appraise Online Health Information. Journal of Pediatrics, 221, 215–223.e215. https://doi.org/10.1016/j.jpeds.2020.02.074

Gandhi, S., Chiu, M., Lam, K., Cairney, J. C., Guttmann, A., & Kurdyak, P. (2016, Feb). Mental Health Service Use Among Children and Youth in Ontario: Population-Based Trends Over Time. Canadian Journal of Psychiatry - Revue Canadienne de Psychiatrie, 61(2), 119–124. https://doi.org/10.1177/0706743715621254

Graham, P., Evitts, T., & Thomas-MacLean, R. (2008, Jul). Environmental scans: how useful are they for primary care research? Canadian Family Physician, 54(7), 1022–1023.

Grist, R., Cliffe, B., Denne, M., Croker, A., & Stallard, P. (2018, Jul). An online survey of young adolescent girls’ use of the internet and smartphone apps for mental health support. BJPsych Open, 4(4), 302–306. https://doi.org/https://dx.doi.org/10.1192/bjo.2018.43

Haavik, L., Joa, I., Hatloy, K., Stain, H. J., & Langeveld, J. (2019, Oct). Help seeking for mental health problems in an adolescent population: the effect of gender. Journal of Mental Health, 28(5), 467–474. https://doi.org/https://dx.doi.org/10.1080/09638237.2017.1340630

Havinga, P. J., Hartman, C. A., Visser, E., Nauta, M. H., Penninx, B., Boschloo, L., & Schoevers, R. A. (2018, 02). Offspring of depressed and anxious patients: Help-seeking after first onset of a mood and/or anxiety disorder [Research Support, Non-U.S. Gov’t]. Journal of Affective Disorders, 227, 618–626. https://doi.org/https://dx.doi.org/10.1016/j.jad.2017.11.017

Headspace. (2021). what is anxiety & the effects on mental health. Retrieved March 28, 2021 from https://headspace.org.au/young-people/what-is-anxiety-and-the-effects-on-mental-health/

Health, F. (2020). 10 Common Symptoms of Anxiety Disorder. Retrieved April 2, 2021 from https://facty.com/conditions/anxiety-disorder/10-common-symptoms-of-anxiety-disorder/?style=quick&utm_source=adwords-

Health, J. C. F. S. (2021). Youth Engagement Toolkit. Retrieved April 21,2021 from http://www.jcsh-cces.ca/explore-resources/youth-engagement/

Health, M. (2020). Online Psychological Therapy. Retrieved March 21, 2021 from http://www.mavenhealth.com/anxiety/index.html

Health, T. M. (2021). Generalized Anxiety Disorder. Retrieved March 27, 2021 from http://mentalhealthliteracy.org/mental-disorders/generalized-anxiety-disorder/

Health, V. B. (2021). Causes, Symptoms & Effects of Anxiety. Retrieved March 27, 2021 from https://www.villagebh.com/disorders/anxiety/symptoms-signs-effects/

Healthdirect. (2020). Anxiety in teenagers. Retrieved March 27, 2021 from https://www.healthdirect.gov.au/anxiety-in-teenagers

healthline. (2021). Anxiety Diagnosis. Retrieved April 2, 2021 from https://www.healthline.com/health/anxiety-diagnosis#outlook

healthychildren.org. (2021). Anxiety in Teens in Rising: What’s Going On? Retrieved April 2, 2021 from https://www.healthychildren.org/English/health-issues/conditions/emotional-problems/Pages/Anxiety-Disorders.aspx

Help, B. (2021). Anxiety Articles. Retrieved April 2, 2021 from https://www.betterhelp.com/advice/anxiety/

HereToHelp. *Online Screenings*. Retrieved April 4, 2021 from https://www.heretohelp.bc.ca/screening/online/?screen=anxiety

HeretoHelp. (2019). For Youth: Learn about Anxiety. Retrieved March 28, 2021 from https://www.heretohelp.bc.ca/infosheet/for-youth-learn-about-anxiety

Hoge, E. A., Ivkovic, A., & Fricchione, G. L. (2012, Nov 27). Generalized anxiety disorder: diagnosis and treatment. BMJ, 345, e7500. https://doi.org/10.1136/bmj.e7500

Hospital, B. C. s. (2021). Anxiety Disorders Symptoms & Causes. Retrieved April 2, 2021 from https://www.childrenshospital.org/conditions-and-treatments/conditions/a/anxiety-disorders/symptoms-and-causes

Hsieh, H.-F., & Shannon, S. E. (2005). Three Approaches to Qualitative Content Analysis. Qualitative Health Research, 15(9), 1277–1288. https://doi.org/10.1177/1049732305276687

inform, N. (2020). Anxiety Disorders in Children. Retrieved April 2, 2021 from https://www.nhsinform.scot/illnesses-and-conditions/mental-health/anxiety-disorders-in-children

Information, M. (2018). Take Control of Your Mental Health. Retrieved April 4, 2021 from https://whatsmym3.com/

InnerHour. The InnerHour Experience. Retrieved April 4, 2021 from https://www.theinnerhour.com/

Institute, B. D. (2021). *Anxiety Self Test*. Retrieved April 4, 2021 from https://www.blackdoginstitute.org.au/resources-support/digital-tools-apps/anxiety-self-test/

Institute, S. F. (2021). Defeat Anxiety. Retrieved March 27, 2021 from https://strongestfamilies.com/defeat-anxiety/

Johnsoni, J. (2021). Global market share of search engines 2010-2021. Retrieved May 10, 2021 from https://www.statista.com/statistics/216573/worldwide-market-share-of-search-engines/

Kendall PC, M. H., Swan A, Carper MM, Mercado R, Kagan E, Crawford E. (2016). What Steps to Take? How to Approach Concerning Anxiety in Youth. Clinical Psychology: Science and Practice, 23(3), 211–229.

Kendall, P. C., Safford, S., Flannery-Schroeder, E., & Webb, A. (2004, Apr). Child anxiety treatment: outcomes in adolescence and impact on substance use and depression at 7.4- year follow-up. Journal of Consulting & Clinical Psychology, 72(2), 276–287. https://doi.org/10.1037/0022-006x.72.2.276

Kenny, R., Dooley, B., & Fitzgerald, A. (2016, Jun). Developing mental health mobile apps: Exploring adolescents’ perspectives. Health Informatics J, 22(2), 265–275. https://doi.org/10.1177/1460458214555041

KidsHealth. (2021). Anxiety. Retrieved March 27, 2021 from https://www.kidshealth.org.nz/anxiety

Lal, S., Nguyen, V., & Theriault, J. (2018, Jun). Seeking mental health information and support online: experiences and perspectives of young people receiving treatment for first-episode psychosis. Early intervention in psychiatry, 12(3), 324–330. https://doi.org/10.1111/eip.12317

Langley, E. L., Wootton, B. M., & Grieve, R. (2018, Aug). The utility of the health belief model variables in predicting help-seeking intention for anxiety disorders [Empirical Study; Quantitative Study]. Australian Psychologist, 53(4), 291–301. https://doi.org/http://dx.doi.org/10.1111/ap.12334

Leech, T., Dorstyn, D. S., & Li, W. (2019, Dec 16). eMental health service use among Australian youth: a cross-sectional survey framed by Andersen. Australian Health Review, 16, 16. https://doi.org/https://dx.doi.org/10.1071/AH19095

LLC, I. H. (2020). *Anxiety Test*. Retrieved April 4, 2021 from https://play.google.com/store/apps/details?id=com.feartools.anxietytest&hl=en_CA&gl=US

LLC, I. H. (December 2016). FearTools. Retrieved April 4 from https://www.feartools.com/

Manafo, E., Petermann, L., Mason-Lai, P., & Vandall-Walker, V. (2018, Feb 7). Patient engagement in Canada: a scoping review of the ’how’ and ’what’ of patient engagement in health research. Health Res Policy Syst, 16(1), 5. https://doi.org/10.1186/s12961-018-0282-4

Manitoba, H. C. Teen Clinic Mental Health Toolkit. Retrieved March 28, 2021 from https://www.gov.mb.ca/healthychild/mcad/teen_clinic_mental_health_toolkit.pdf

Manual, M. (2019). *Generalized Anxiety Disorder in Children*. Retrieved March 27, 2021 from https://www.merckmanuals.com/home/children-s-health-issues/mental-health-disorders-in-children-and-adolescents/generalized-anxiety-disorder-in-children

McRae, L., O’Donnell, S., Loukine, L., Rancourt, N., & Pelletier, C. (2016a, Dec). Report summary - Mood and Anxiety Disorders in Canada, 2016. Health Promotion and Chronic Disease Prevention in Canada, 36(12), 314–315. https://doi.org/10.24095/hpcdp.36.12.05 (Note de synthèse - Les troubles anxieux et de l’humeur au Canada, 2016.)

McRae, L., O’Donnell, S., Loukine, L., Rancourt, N., & Pelletier, C. (2016b). Report summary - Mood and Anxiety Disorders in Canada, 2016 [Note de synthèse - Les troubles anxieux et de l’humeur au Canada, 2016]. Health promotion and chronic disease prevention in Canada : research, policy and practice, 36(12), 314–315. https://doi.org/10.24095/hpcdp.36.12.05

Mind, M. y. (2021). Generalized Anxiety Disorder. Retrieved March 24 from https://mindyourmind.ca/illnesses/generalized-anxiety-disorder-0

Minds, Y. (2021). Anxiety. Retrieved April 4 from https://youngminds.org.uk/find-help/conditions/anxiety/

Moir, F., Fernando, A. T., 3rd, Kumar, S., Henning, M., Moyes, S. A., & Elley, C. R. (2015, Mar 27). Computer Assisted Learning for the Mind (CALM): the mental health of medical students and their use of a self-help website [Research Support, Non-U.S. Gov’t]. New Zealand Medical Journal, 128(1411), 51–58. https://resolver.library.ualberta.ca/resolver?sid=OVID:medline&id=pmid:25820503&id=doi:&issn=0028-8446&isbn=&volume=128&issue=1411&spage=51&pages=51-8&date=2015&title=New+Zealand+Medical+Journal&atitle=Computer+Assisted+Learning+for+the+Mind+%28CALM%29%3A+the+mental+health+of+medical+students+and+their+use+of+a+self-help+website.&aulast=Moir&pid=%3Cauthor%3EMoir+F%2CFernando+AT+3rd%2CKumar+S%2CHenning+M%2CMoyes+SA%2CElley+CR%3C%2Fauthor%3E%3CAN%3E25820503%3C%2FAN%3E%3CDT%3EJournal+Article%3C%2FDT%3E

Narmandakh, A., Roest, A. M., Jonge, P., & Oldehinkel, A. J. (2020, 03). The bidirectional association between sleep problems and anxiety symptoms in adolescents: a TRAILS report [Research Support, Non-U.S. Gov’t]. Sleep Medicine, 67, 39–46. https://doi.org/https://dx.doi.org/10.1016/j.sleep.2019.10.018

Newman, M. G., Zuellig, A. R., Kachin, K. E., Constantino, M. J., Przeworski, A., Erickson, T., & Cashman-McGrath, L. (2002, 2002/03/01/). Preliminary reliability and validity of the generalized anxiety disorder questionnaire-IV: A revised self-report diagnostic measure of generalized anxiety disorder. Behavior Therapy, 33(2), 215–233. https://doi.org/https://doi.org/10.1016/S0005-7894(02)80026-0

NHS. (2020a). Anxiety disorders in children. Retrieved March 27, 2021 from https://www.nhs.uk/mental-health/children-and-young-adults/advice-for-parents/anxiety-disorders-in-children/

NHS. (2020b). Depression and anxiety self-assessment quiz. Retrieved April 4, 2021 from https://www.nhs.uk/mental-health/self-help/guides-tools-and-activities/depression-anxiety-self-assessment-quiz/

Nutt, D., Argyropoulos, S., Hood, S., & Potokar, J. (2006, Jul). Generalized anxiety disorder: A comorbid disease. European Neuropsychopharmacology, 16 Suppl 2, S109–118. https://doi.org/10.1016/j.euroneuro.2006.04.003

Ontario, A. D. A. o. (2021). Anxiety and Youth. Retrieved March 27, 2021 from http://www.anxietydisordersontario.ca/anxiety-resource-centre/anxiety-and-youth/

Ontario, M. D. A. o. Frequently Asked Questions - Anxiety and Mood Disorders. Retrieved March 27, 2021 from https://mooddisorders.ca/faq/anxiety-and-mood-disorders

Pelletier, L., O’Donnell, S., McRae, L., & Grenier, J. (2017, Feb). The burden of generalized anxiety disorder in Canada. Health Promotion and Chronic Disease Prevention in Canada, 37(2), 54–62. https://doi.org/10.24095/hpcdp.37.2.04 (Le fardeau du trouble d’anxiété généralisée au Canada.)

Phillips, S. P., & Yu, J. (2021, Mar 1). Is anxiety/depression increasing among 5-25 year-olds? A cross-sectional prevalence study in Ontario, Canada, 1997-2017. Journal of Affective Disorders, 282, 141–146. https://doi.org/10.1016/j.jad.2020.12.178

Phone, K. H. Questionnaire: Reflecting on feelings of anxiety. Retrieved April 4, 2021 from https://kidshelpphone.ca/get-info/questionnaire-reflecting-on-feelings-of-anxiety/

Pine, D. S., Cohen, P., Gurley, D., Brook, J., & Ma, Y. (1998, Jan). The risk for early-adulthood anxiety and depressive disorders in adolescents with anxiety and depressive disorders. Archives of General Psychiatry, 55(1), 56–64. https://doi.org/10.1001/archpsyc.55.1.56

Pittsburgh, U. o. (2021). Child and Adolescent Bipolar Spectrum Services. Retrieved April 4, 2021 from https://www.pediatricbipolar.pitt.edu/resources/instruments

Pretorius, C., Chambers, D., Cowan, B., & Coyle, D. (2019, Aug 26). Young People Seeking Help Online for Mental Health: Cross-Sectional Survey Study. JMIR Mental Health, 6(8), e13524. https://doi.org/10.2196/13524

Prevention, C. f. D. C. a. (2020). Anxiety and Depression in Children. Retrieved March 28, 2021 from https://www.cdc.gov/childrensmentalhealth/depression.html

Prospects, W. E. S. a. (2014). Country classification. Retrieved April 21, 2021 from https://www.un.org/en/development/desa/policy/wesp/wesp_current/2014wesp_country_ classification.pdf

PsychCentral. *Anxiety Screening Test*. Retrieved April 4, 2021 from https://psychcentral.com/quizzes/anxiety-quiz#Learn-More-About-Anxiety

Psychiatry, A. A. o. C. A. *Your Adolescent - Anxiety and Avoidant Disorders*. Retrieved April 2, 2021 from https://www.aacap.org/AACAP/Families_and_Youth/Resource_Centers/Anxiety_Disorder_Resource_Center/Your_Adolescent_Anxiety_and_Avoidant_Disorders.aspx

Psycom. (2021a). 6 Hidden Signs of Teen Anxiety. Retrieved April 2, 2021 from https://www.psycom.net/hidden-signs-teen-anxiety/

PSYCOM. (2021b). Anxiety Test (Self-Assessment). Retrieved April 4, 2021 from https://www.psycom.net/anxiety-test

Purcell, R., Jorm, A. F., Hickie, I. B., Yung, A. R., Pantelis, C., Amminger, G. P., Glozier, N., Killackey, E., Phillips, L. J., Wood, S. J., Harrigan, S., Mackinnon, A., Scott, E., Hermens, D. F., Guastella, A. J., Kenyon, A., Mundy, L., Nichles, A., Scaffidi, A., Spiliotacopoulos, D., Taylor, L., Tong, J. P., Wiltink, S., Zmicerevska, N., & McGorry, P. D. (2015, Dec). Demographic and clinical characteristics of young people seeking help at youth mental health services: baseline findings of the Transitions Study [Research Support, Non-U.S. Gov’t]. Early intervention in psychiatry, 9(6), 487–497. https://doi.org/https://dx.doi.org/10.1111/eip.12133

raisingchildren.net.au. (2021). Anxiety disorders in teenagers. Retrieved April 2, 2021 from https://raisingchildren.net.au/pre-teens/mental-health-physical-health/stress-anxiety-depression/anxiety-disorders

Research, C. I. o. H. (2012). Canada’s strategy for patient-oriented research: improving health outcomes through evidence-informed care. Retrieved April 19, 2021 from https://cihr-irsc.gc.ca/e/44000.html

Research, C. I. o. H. (2019). Strategy for patient-oriented research. www.cihr-irsc.gc.ca/e/41204.html

Rickwood, D. J., Deane, F. P., & Wilson, C. J. (2007, Oct 1). When and how do young people seek professional help for mental health problems? Medical Journal of Australia, 187(S7), S35–39. https://doi.org/10.5694/j.1326-5377.2007.tb01334.x

Romanson, E. E. (2018). Online help seeking in emerging adults: The role of attachment style, emotion regulation, and distress disclosure [Dissertation Empirical Study; Quantitative Study]. Dissertation Abstracts International: Section B: The Sciences and Engineering, 79(10-B(E)), No Pagination Specified. https://resolver.library.ualberta.ca/resolver?sid=OVID:psycdb&id=pmid:&id=doi:&issn=0419-4217&isbn=978-0355968484&volume=79&issue=10-B%28E%29&spage=No&pages=No+Pagination+Specified&date=2018&title=Dissertation+Abstracts+International%3A+Section+B%3A+The+Sciences+and+Engineering&atitle=Online+help+seeking+in+emerging+adults%3A+The+role+of+attachment+style%2C+emotion+regulation%2C+and+distress+disclosure.&aulast=Romanson&pid=%3Cauthor%3ERomanson%2C+Emily+E.+J%3C%2Fauthor%3E%3CAN%3E2018-34221-143%3C%2FAN%3E%3CDT%3EDissertation%3C%2FDT%3E

Scrandis, D. A. (2019). Anxiety disorders in adolescents. Nurse Practitioner, 44(8), 12–14. https://doi.org/10.1097/01.NPR.0000559851.14367.c6

service, o. h. (2020). Anxiety disorders in children. Retrieved March 27, 2021 from https://www2.hse.ie/conditions/mental-health/anxiety-disorders-in-children.html

Sigmund, H. (2021). Anxiety in Teens - How to Help a Teenager Deal with Anxiety. Retrieved April 2, 2021 from https://www.heysigmund.com/anxiety-in-teens/

Sinai, C. (2021). Generalized Anxiety Disorder (GAD) in Children and Teens. Retrieved April 2, 2021 from https://www.cedars-sinai.org/health-library/diseases-and-conditions---pediatrics/g/generalized-anxiety-disorder-gad-in-children.html

Society, T. C. s. *Anxiety*. Retrieved March 27, 2021 from https://www.childrenssociety.org.uk/information/young-people/well-being/resources/anxiety

Solutions, B. S. (2017). WellMind. Retrieved April 4, 2021 from https://play.google.com/store/apps/details?id=com.bluestepsolutions.wellmind&hl=en_CA&gl=US

Spence, S. H., Zubrick, S. R., & Lawrence, D. (2018, May). A profile of social, separation and generalized anxiety disorders in an Australian nationally representative sample of children and adolescents: Prevalence, comorbidity and correlates. Australian & New Zealand Journal of Psychiatry, 52(5), 446–460. https://doi.org/10.1177/0004867417741981

Spitzer, R. L., Kroenke, K., Williams, J. B. W., & Löwe, B. (2006). A Brief Measure for Assessing Generalized Anxiety Disorder: The GAD-7. Archives of Internal Medicine, 166(10), 1092–1097. https://doi.org/10.1001/archinte.166.10.1092

Stem4. (2012). Anxiety. Retrieved March 28, 2021 from https://stem4.org.uk/anxiety/

Substance, A., & Mental Health Services, A. (2016). CBHSQ Methodology Report. In Impact of the DSM-IV to DSM-5 Changes on the National Survey on Drug Use and Health. Substance Abuse and Mental Health Services Administration (US).

Summerhurst, C., Wammes, M., Wrath, A., & Osuch, E. (2017, 01). Youth Perspectives on the Mental Health Treatment Process: What Helps, What Hinders? [Research Support, Non-U.S. Gov’t]. Community Mental Health Journal, 53(1), 72–78. https://doi.org/https://dx.doi.org/10.1007/s10597-016-0014-6

Taylor, J. (2015). Pocket Mood Tracker. Retrieved April 4, 2021 from https://apps.apple.com/us/app/pocket-mood-tracker/id960876692

TeensHealth. (2014). Anxiety Disorders. Retrieved March 28, 2021 from https://kidshealth.org/en/teens/anxiety.html?WT.ac=ctg#catmental-health

Today, P. (2021). Anxiety Test. Retrieved April 4, 2021 from https://www.psychologytoday.com/ca/tests/health/anxiety-test

Tricco, A. C., Lillie, E., Zarin, W., O’Brien, K. K., Colquhoun, H., Levac, D., Moher, D., Peters, M. D. J., Horsley, T., Weeks, L., Hempel, S., Akl, E. A., Chang, C., McGowan, J., Stewart, L., Hartling, L., Aldcroft, A., Wilson, M. G., Garritty, C., Lewin, S., Godfrey, C. M., Macdonald, M. T., Langlois, E. V., Soares-Weiser, K., Moriarty, J., Clifford, T., Tunçalp, Ö., & Straus, S. E. (2018, 2018/10//). PRISMA Extension for Scoping Reviews (PRISMA-ScR): Checklist and Explanation. Annals of Internal Medicine, 169(7), 467–473. https://doi.org/10.7326/m18-0850

Understood. (2021). Signs of Anxiety in Tweens and Teens. Retrieved April 2, 2021 from https://www.understood.org/en/friends-feelings/managing-feelings/stress-anxiety/signs-your-teen-or-tween-is-struggling-with-anxiety

Van Ameringen, M., Simpson, W., Patterson, B., & Turna, J. (2015, Dec 15). Internet screening for anxiety disorders: Treatment-seeking outcomes in a three-month follow-up study. Psychiatry Research, 230(2), 689–694. https://doi.org/https://dx.doi.org/10.1016/j.psychres.2015.10.031

Wehry, A. M., Beesdo-Baum, K., Hennelly, M. M., Connolly, S. D., & Strawn, J. R. (2015, Jul). Assessment and treatment of anxiety disorders in children and adolescents. Current Psychiatry Reports, 17(7), 52. https://doi.org/10.1007/s11920-015-0591-z

Wilburn, A., Vanderpool, R. C., & Knight, J. R. (2016, Aug 18). Environmental Scanning as a Public Health Tool: Kentucky’s Human Papillomavirus Vaccination Project. Prev Chronic Dis, 13, E109. https://doi.org/10.5888/pcd13.160165

