## Supplemental Table 1 for "Online anxiety resources for Canadian youth: a systematic environmental scan"

**Supplementary Table 1: Characteristics of Information Sources Describing Signs and Symptoms of Anxiety**

| **First Author (Country)**  **Study Design (References)**  **Organization Type** | **Condition**  **Symptoms are Listed**  **Audience (i.e, Youth, Parents)** | **Objective of Resource**  **Business looking to Attract Clients** | **Next Steps Provided**  **Linked to a Self-Screening Resource** |
| --- | --- | --- | --- |
| Anxiety Disorders Centre (Canada)^1^  Website (no references)  Private - individual/company | GAD  Yes  NA | Information about various conditions with intent to contact psychological therapy  Yes | Yes; organizations’ psychological services  No |
| Link: <http://www.mavenhealth.com/anxiety/index.html> | | | |
| Mind your Mind (Canada)^2^  Website (no references)  Federal Government | GAD  Yes  Young people aged 14 to 29 | Describe GAD  No | Yes; link to help section on website  No |
| Link: <https://mindyourmind.ca/illnesses/generalized-anxiety-disorder-0> | | | |
| Mood Disorders Association of Ontario (Canada)^3^  Website (descriptive with references)  Private – foundation/charity | Various and GAD  No  NA | FAQ about anxiety and mood disorders  No | Yes; links to "Where else can I go to learn more about anxiety and mood disorders?"  No |
| Link: <https://mooddisorders.ca/faq/anxiety-and-mood-disorders> | | | |
| Strongest Families Institute (Canada)^4^  Website (no references)  Private - foundation/charity | Anxiety disorders  Yes  Youth 12-17 years | To provide information about the mental health services their charity provides  No | Yes; need referral from local health providers to access free services  No |
| Link: <https://strongestfamilies.com/defeat-anxiety/> | | | |
| Youth in BC (Canada)^5^  Website (no references)  Provincial Government | GAD  Yes  Youth in BC | Resource and chat for youth experiencing a variety of issues  No | Yes; chat line, links to external sites and provincial services  No |
| Link: <https://youthinbc.com/youth-issues-2/mental-health/anxiety/> | | | |
| Barzilay (USA)^6^  Cross-sectional  Academic | GAD  No  Youth 11-21 years | Characterize anxiety symptoms suggestive of risk for depression and suicidal ideation (SI) in community youths  No | No  No |
| Link: NA | | | |
| Narmandakh (Australia)^7^  Prospective Cohort  Academic | Anxiety Disorders  No  Youth 11-25 years | Longitudinal study aimed to examine bidirectional associations between sleep problems and anxiety symptoms  No | No  No |
| Link: NA | | | |
| Farmer (USA)^8^  Qualitative  Academic | Anxiety Disorders  No  Youth 18-23 years | Qualitatively examine career anxiety; the experience of anxiety that is embedded in the career concerns of college students as they engage in the career development process  No | No  No |
| Link: NA | | | |
| Wehry (USA)^9^  Narrative review  Academic | Anxiety Disorders  Yes  Children and adolescents | The article reviews the current epidemiology, longitudinal trajectory, and neurobiology of anxiety disorders in youth, and treatments  No | No  No |
| Link: NA | | | |
| Kendall (USA)^10^  Narrative review  Academic | Anxiety Disorders  No  Youth | Outlines steps to take for youth with anxiety  No | No  No |
| Link: NA | | | |
| Arcaro (Canada)^11^  Cross-sectional  Academic | Anxiety and Mood Disorders  Yes  Youth 16-26 years | Investigate presenting concerns of emerging adults when seeking treatment at an early intervention program for anxiety disorders.  No | Yes; refers user to First Episode Mood and Anxiety Program (FEMAP) in London, Ontario  No |
| Link: NA | | | |
| Bandelow (Germany)^12^  Narrative review  Academic | Anxiety Disorders  No  NA | Outlines the epidemiology of anxiety disorders  No | No  No |
| Link: NA | | | |
| CMHA: Children, Youth, and Anxiety (Canada)^13^  Website (no references)  Federal Government | Anxiety Disorders  Yes  Children and youth | Describing anxiety problems and signs  No | Yes; provides link to internal CMHA locations to contact  No |
| Link: https://cmha.ca/documents/children-youth-and-anxiety | | | |
| Anxiety Disorders Association of Ontario (Canada)^14^  Website (with links and referneces)  Provincial Government | Anxiety Disorders  Yes  Youth and parents | Description and resourced to help children and teenagers living with anxiety  No | Yes; provides links to local emergency departments, crisis lines, walk-in counselling, support and self-help groups  No |
| Link: http://www.anxietydisordersontario.ca/anxiety-resource-centre/anxiety-and-youth/ | | | |
| Anxiety Centre (Canada)^15^  Website (references at end)  Private - individual/company | Anxiety Disorders  Yes  NA | Frequent Questions about Anxiety, Anxiety Disorder, Anxiety Attacks  No | Yes; short section on anxiety disorders with links to references  No |
| Link: https://www.anxietycentre.com/anxiety-faq.shtml | | | |
| Teen Mental Health.org (Canada)^16^  Website (no references)  Private - individual/company | GAD  Yes  Youth (teenagers) | Description and symptoms of GAD  No | Yes; links to external organizations  No |
| Link: http://teenmentalhealth.org/mental-disorders/generalized-anxiety-disorder/ | | | |
| InnerHour:Self-Care Therapy (India)^17^  Website (business no references)  Private - individual/company | Anxiety Disorders  No  NA | Describe how services can help with anxiety  Yes | Yes: their online app to manage various disorders  No |
| Link: https://www.theinnerhour.com/anxiety | | | |
| WellMind (UK)^18^  App (no references)  Private - individual/company | Anxiety Disorders  No  NA | To help with managing anxiety  No | Yes; helpline numbers and NHS resources  No |
| Link: https://play.google.com/store/apps/details?id=com.bluestepsolutions.wellmind&hl=en_CA | | | |
| Village Behavioral Health (USA)^19^  Website (no references)  Private - foundation/charity | GAD  Yes  Youth 13-17 | State Rehab Program for youth; Causes, Symptoms and Effects of Anxiety  No | Yes; possible admission to program  No |
| Link: https://www.villagebh.com/disorders/anxiety/symptoms-signs-effects/ | | | |
| Merck Manual (USA)^20^  Website (no references)  Private - individual/company | GAD  Yes  Children | Describe GAD symptoms, diagnosis, treatments  No | No  No |
| Link: <https://www.merckmanuals.com/home/children-s-health-issues/mental-health-disorders-in-children-and-adolescents/generalized-anxiety-disorder-in-children> | | | |
| Healthdirect (Australia)^21^  Website (sources listed at end)  Federal Government | Anxiety Disorders  Yes  Youth (teenagers) | To describe anxiety in teenagers  No | Yes; where to get help section; doctors, local programs  No |
| Link: https://www.healthdirect.gov.au/anxiety-in-teenagers | | | |
| OurHealthService (UK)^22^  Website (no references)  Federal Government | Anxiety Disorders  Yes  Children and parents | To describe symptoms, treatments, where to get help, causes  No | Yes; help phone line  No |
| Link: https://www2.hse.ie/conditions/mental-health/anxiety-disorders-in-children.html | | | |
| KidsHealth (New Zealand)^23^  Website (few references at end)  Private - foundation/charity | Anxiety Disorders  No  Children | Provide a general description of anxiety, treatments, causes, and different types  No | Yes; help phone and links to local health agencies  No |
| Link: https://www.kidshealth.org.nz/anxiety | | | |
| Aspiro (USA)^24^  Website (no references)  Private - individual/company | Anxiety Disorders  Yes  Youth (young adults and teenagers) | Describes the signs of anxiety in young adults and teenagers  Yes | Yes; wilderness adventure  No |
| Link: https://aspiroadventure.com/blog/13-signs-of-anxiety-in-teens-and-young-adults/ | | | |
| NHS: Anxiety disorders in children (UK)^25^  Website (few links to other sites)  Federal Government | Anxiety Disorders  Yes  Children and parents | Describes anxiety disorders in children  No | Yes; help phone and links to local health agencies  No |
| Link: https://www.nhs.uk/conditions/anxiety-disorders-in-children/ | | | |
| The Children's Society (UK)^26^  Website (no references)  Private - foundation/charity | Anxiety Disorders  Yes  Children | Describes anxiety disorders in children  No | Yes; links to top applications for anxiety (none are assessments)  No |
| Link: https://www.childrenssociety.org.uk/information/young-people/well-being/resources/anxiety | | | |
| Family Centre (Canada)^27^  Website (no references)  Private - foundation/charity | Anxiety Disorders  Yes  Children and parents | Describes anxiety signs/symptoms in children  No | Yes; local phone support number but link for more information does not work  No |
| Link: https://www.famcentre.ca/anxiety-in-children-and-youth/ | | | |
| Mind Diagnostics (USA)^28^  Website (no references)  Private - individual/company | Anxiety Disorders  Yes  NA | Describe anxiety and provides links to their counseling services  Yes | Yes; link to application or online test; upon results of online test given contacts for online counseling  Yes |
| Link: https://www.mind-diagnostics.org/anxiety-test?utm_source=AdWords&utm_medium=Search_PPC_c&utm_term=anxiety%20disorder_b&utm_content=88328057960&network=g&placement=&target=&matchtype=b&utm_campaign=6453264419&ad_type=mind-diagnostics&adposition=&gclid=Cj0K | | | |
| Scrandis (USA)^29^  Descriptive  Academic | Anxiety Disorders  Yes  Youth (adolescents) | Description of anxiety disorders in adolescents  No | No  No |
| Link: NA | | | |
| Spence (Australia)^30^  Cross-sectional  Academic | GAD  No  Youth 4-17 years | Describes 12-month prevalence of social anxiety disorder, separation anxiety disorder, and GAD in a sample of Australian youth  No | No  No |
| Link: NA | | | |
| Information about Mood & Anxiety (Canada )^31^  Website (no references)  Private – individual/company | GAD  Yes  Youth | Describing anxiety briefly in the context of explaining applications created to treat it  No | No  No |
| Link: https://www.lhsc.on.ca/femap-first-episode-mood-and-anxiety-program/information-about-mood-anxiety | | | |
| Anxious children: Treating a crippling disorder (Canada) ^32^  Website (no references)  Academic | Anxiety  Yes  Children and Parents | Describes the treatment of anxiety and provides a background on the signs and symptoms  No | Yes; provides a crisis number and promotes help-seeking  No |
| Link: https://www.thechildren.com/health-info/conditions-and-illnesses/anxious-children-treating-crippling-disorder | | | |
| Child and Youth Mental Health (Canada)^33^  Website (no references)  Provincial Government | Anxiety Disorders  Yes  Youth | Understanding anxiety  No | Yes; promotes help-seeking, provides kids help phone number and other links  No |
| Link: https://ontario.cmha.ca/mental-health/child-and-youth-mental-health/ | | | |
| Anxiety Disorders (Canada)^34^  Website (no references)  Provincial Government | Anxiety Disorders  Yes  NA | Describes anxiety and signs and symptoms  No | Yes; provides various options for treatment, links, phone numbers  No |
| Link: https://cmha.bc.ca/documents/anxiety-disorders/ | | | |
| Anxiety Disorders (USA)^35^  Website (no references)  Private - foundation/charity | Anxiety Disorders  Yes  Youth | Describe anxiety  No | Yes; promotes help-seeking and regular exercise and nutrition. No links or other resources provided  No |
| Link: https://kidshealth.org/en/teens/anxiety.html?WT.ac=ctg#catmental-health | | | |
| Anxiety (Canada)^36^  Website (no references)  Provincial Government | Anxiety  Yes  Youth | Describes anxiety and signs and symptoms  No | Yes; provides tips, apps and tools, promotes help seeking and provides a get support option  No |
| Link: https://foundrybc.ca/articles/anxiety-the-basics/?return_page=1292 | | | |
| For Youth: Learn about Anxiety (Canada)^37^  Website (no references)  Provincial Government | Anxiety  Yes  Youth | Describes signs and symptoms in great detail and providing resources  No | Yes; provides links to more anxiety info and self-help programs  No |
| Link: https://www.heretohelp.bc.ca/infosheet/for-youth-learn-about-anxiety | | | |
| Teen Clinic Mental Health Toolkit (Canada)^38^  PDF with resources (references at end of document)  Provincial Government | Anxiety  Yes  Youth | Document covers a variety of mental health topics and challenges that youth might face; describes anxiety and provides resources  No | Yes; links to additional resources, breathing exercises, distress lines  Yes; embedded anxiety self-assessment |
| Link: https://www.gov.mb.ca/healthychild/mcad/teen_clinic_mental_health_toolkit.pdf | | | |
| Stem4 (Canada)^39^  Website (no references)  Private - foundation/charity | Anxiety  Yes  Youth | Describes anxiety and provides resources  No | Yes; encourages help seeking, and provides an application  No |
| Link: https://stem4.org.uk/anxiety/ | | | |
| Headspace (Australia)^40^  Website (no references)  Federal Government | Anxiety  Yes  Youth | Describes anxiety and provides resources  No | Yes; encourages help-seeking, next steps for managing symptoms, applications for managing anxiety, and professional support contacts  No |
| Link: https://headspace.org.au/young-people/what-is-anxiety-and-the-effects-on-mental-health/ | | | |
| CDC (USA)^41^  Website (no references)  Federal Government | Anxiety and Depression  Yes  Children | Describes anxiety and provides resources  No | Yes; links to more information, contact information for a clinician, and links to preventative care  No |
| Link: https://www.cdc.gov/childrensmentalhealth/depression.html | | | |
| American Academy of Child and Adolescent Psychiatry (USA)^42^  Website (no references)  Federal Government | Anxiety Disorders  Yes  Youth | Describes anxiety and provides resources  No | Yes; includes links to more information on anxiety in children and youth and a resource center  No |
| Link: https://www.aacap.org/AACAP/Families_and_Youth/Resource_Centers/Anxiety_Disorder_Resource_Center/Your_Adolescent_Anxiety_and_Avoidant_Disorders.aspx | | | |
| Healthline (USA)^43^  Website (no references)  Private - individual/company | Anxiety Disorders  Yes  NA | Describes anxiety and next steps  No | Yes; different forms of treatment such as medication, counselling, lifestyle choices, and communication  No |
| Link: https://www.healthline.com/health/anxiety-diagnosis#physical-exam | | | |
| Newport Academy (USA)^44^  Website (descriptive with few embedded references)  Private - individual/company | Anxiety Disorders  Yes  Youth and parents | Describes anxiety and next steps while providing some resources  No | Yes; provides links to more anxiety info but no phone numbers or links to resources  No |
| Link: https://www.newportacademy.com/resources/mental-health/how-to-recognize-teen-anxiety-disorders/ | | | |
| Cleveland Clinic (USA)^45^  Website (no references)  Private - individual/company | Anxiety Disorders  Yes  NA | Describes anxiety and next steps  Yes | Yes; suggests help seeking but does not provide any resources  No |
| Link: https://my.clevelandclinic.org/health/diseases/9536-anxiety-disorders | | | |
| Understood.com (USA)^46^  Website (no references)  Private - foundation/charity | Anxiety Disorders  Yes  Youth | Describes anxiety signs and symptoms  No | Yes; promotes help-seeking and intervention but no new links or resources provided  No |
| Link: https://www.understood.org/en/friends-feelings/managing-feelings/stress-anxiety/signs-your-teen-or-tween-is-struggling-with-anxiety | | | |
| raisingchildren.net.au (Australia)^47^  Website (no references)  Federal Government | Anxiety Disorders  Yes  Youth and parents | Describes anxiety signs and symptoms while providing some resources  No | Yes; extensive list of resources embedded within the main text, including distress line, contact with physician or psychologist  No |
| Link: https://raisingchildren.net.au/pre-teens/mental-health-physical-health/stress-anxiety-depression/anxiety-disorders | | | |
| Boston Children's Hospital (USA)^48^  Website (no references)  Private - individual/company | GAD  Yes  Children | Describes GAD  No | No  No |
| Link: https://www.childrenshospital.org/conditions-and-treatments/conditions/a/anxiety-disorders/symptoms-and-causes | | | |
| Better Help (USA)^49^  Website (embedded references)  Private - individual/company | Anxiety  Yes  NA | Describes anxiety and signs and symptoms  Yes | Yes; provides links to the companies mental health supports (for payment)  No |
| Link: https://www.betterhelp.com/advice/anxiety/how-to-tell-if-you-have-anxiety-10-signs-and-symptoms/?network=g&placement=&target=&matchtype=b&ad_type=text&utm_source=AdWords&utm_medium=Search_PPC_c&utm_term=_b&utm_content=107920132930&network=g&placement=&tar | | | |
| NHS inform (Scotland)^50^  Website (no references)  Federal Government | Anxiety  Yes  Children | Describes anxiety and signs and symptoms. Helps identify next steps.  No | Yes; promotes help-seeking from a doctor or other organizations, links to explore youth counselling  No |
| Link: https://www.nhsinform.scot/illnesses-and-conditions/mental-health/anxiety-disorders-in-children | | | |
| Cedars Sinai (USA)^51^  Website (no references)  Private - foundation/charity | GAD  Yes  Youth and parents | Describes anxiety and signs and symptoms. Helps identify next steps.  No | Yes; promotes help-seeking but does not provide resources  No |
| Link: https://www.cedars-sinai.org/health-library/diseases-and-conditions---pediatrics/g/generalized-anxiety-disorder-gad-in-children.html | | | |
| Facty Health (Canada)^52^  Website (no references)  Private - individual/company | Anxiety  Yes  NA | Describes anxiety and signs and symptoms.  No | No  No |
| Link: https://facty.com/conditions/anxiety-disorder/10-common-symptoms-of-anxiety-disorder/?style=quick&utm_source=adwords-ca&adid=413214662072&ad_group_id=89271669245&utm_medium=c-search&utm_term=%2Banxiety&utm_campaign=FH-CA---Search---Anxiety-Symptoms---Desk | | | |
| Brain and Behavior (USA)^53^  Website (no references)  Private - foundation/charity | Anxiety  No  NA | Describes anxiety and how it is treated.  No | Yes; links to an anxiety webinar  No |
| Link: https://www.bbrfoundation.org/faq/frequently-asked-questions-about-anxiety?gclid=CjwKCAiAnvj9BRA4EiwAuUMDf4uY-2P1fnBqaJJ29vn-VNgfeO3w7zV68OWpkgA0AXqZZjUGjGw-yhoCm1EQAvD_BwE | | | |
| Psycom (USA)^54^  Website (no references)  Private - individual/company | Anxiety  Yes  Youth | Describes anxiety and signs and symptoms.  No | Yes; promotes help-seeking but does not provide resources  No |
| Link: https://www.psycom.net/hidden-signs-teen-anxiety/ | | | |
| Healthychildren.org (USA)^55^  Website (no references)  Private - foundation/charity | Anxiety  Yes  Youth | Describes anxiety in youth  No | Yes; promotes help-seeking, provides access to an anxiety resource center, and links other articles about anxiety  No |
| Link: https://www.healthychildren.org/English/health-issues/conditions/emotional-problems/Pages/Anxiety-Disorders.aspx | | | |
| Hey Sigmund (Australia)^56^  Website (no references)  Private - individual/company | Anxiety  Yes  Parents | Describes anxiety in youth  No | Yes; provides applications to help manage anxiety and other article links  No |
| Link: https://www.heysigmund.com/anxiety-in-teens/ | | | |
| University of Rochester Medical Center (USA)^57^  Website (no references)  Private - individual/company | GAD  Yes  Youth | Describes anxiety in youth and children  No | Yes; promotes help-seeking and provides links to contact mental health professionals, hospitals to make an appointment, and anxiety information websites  No |
| Link: https://www.urmc.rochester.edu/encyclopedia/content.aspx?ContentTypeID=90&ContentID=P02565 | | | |
| ScreenAgers (USA)^58^  Website (no references)  Private - individual/company | Anxiety  No  Youth and parents | Describes anxiety in youth and children  No | Yes; provides information for different organizations providing counselling support, and phone and text numbers for mental health support  No |
| Link: https://www.screenagersmovie.com/tech-talk-tuesdays/anxiety-what-every-young-person-should-know | | | |
| Young Minds (UK)^59^  Website (no references)  Private - foundation/charity | Anxiety  Yes  Youth and children | Describes anxiety in youth and children  No | Yes; panic line for children (<19), crisis messenger, and website links about medications to treat anxiety, and counselling / therapy  No |
| Link: https://youngminds.org.uk/find-help/conditions/anxiety/ | | | |
| Evolve Treatment (USA)^60^  Website (no references)  Private - individual/company | Anxiety  Yes  Youth (teenagers) and parents | Describes anxiety in teens  Yes | Yes; provides options to explore. Suggests talking to your teenager, findings treatment for them, and exploring medication. Then provides a phone number for a consultation with one of their counsellors  No |
| Link: https://evolvetreatment.com/for-parents/parent-guides/generalized-anxiety-disorder/ | | | |
| MACSCREEN (Canada)^61^  Website (no references)  Provincial Government | GAD  Yes  NA | Describes facts, causes, and treatments of GAD  No | No  No |
| Link: https://www.macanxiety.com/information-about-anxiety-disorders/generalized-anxiety-disorder/ | | | |
| CALM Website (New Zealand)^62^  Website (links to local organizations)  Academic | Anxiety  Yes  NA | General description of anxiety  No | Yes; provides phone numbers for further help from University Health and Counselling Service and links to other anxiety resources  No |
| Link: https://www.calm.auckland.ac.nz/ | | | |

**Abbreviations**: CALM: Computer Assisted Learning for the Mind; GAD = Generalised Anxiety Disorder; NA: Not Available

1.	Health M. Online Psychological Therapy. <http://www.mavenhealth.com/anxiety/index.html>. Published 2020. Accessed March 21, 2021.

2.	Mind My. Generalized Anxiety Disorder. <https://mindyourmind.ca/illnesses/generalized-anxiety-disorder-0>. Published 2021. Accessed March 24, 2021.

3.	Ontario MDAo. Frequently Asked Questions - Anxiety and Mood Disorders. <https://mooddisorders.ca/faq/anxiety-and-mood-disorders>. Accessed March 27, 2021.

4.	Institute SF. Defeat Anxiety. <https://strongestfamilies.com/defeat-anxiety/>. Published 2021. Accessed March 27, 2021.

5.	BC TCIaSPCo. Anxiety. <https://youthinbc.com/youth-issues-2/mental-health/anxiety/>. Published 2013. Accessed March 27, 2021.

6.	Barzilay R, White LK, Moore TM, et al. Association of anxiety phenotypes with risk of depression and suicidal ideation in community youth. *Depress Anxiety.* 2020;05:05.

7.	Narmandakh A, Roest AM, Jonge P, Oldehinkel AJ. The bidirectional association between sleep problems and anxiety symptoms in adolescents: a TRAILS report. *Sleep Med.* 2020;67:39-46.

8.	Farmer C, Thienemann M, Leibold C, Kamalani G, Sauls B, Frankovich J. Psychometric Evaluation of the Caregiver Burden Inventory in Children and Adolescents With PANS. *J Pediatr Psychol.* 2018;43(7):749-757.

9.	Wehry AM, Beesdo-Baum K, Hennelly MM, Connolly SD, Strawn JR. Assessment and treatment of anxiety disorders in children and adolescents. *Curr Psychiatry Rep.* 2015;17(7):52.

10.	Kendall PC MH, Swan A, Carper MM, Mercado R, Kagan E, Crawford E. What Steps to Take? How to Approach Concerning Anxiety in Youth. *CLINICAL PSYCHOLOGY: SCIENCE AND PRACTICE.* 2016;23(3):211 - 229.

11.	Arcaro J, Summerhurst C, Vingilis E, Wammes M, Osuch E. Presenting concerns of emerging adults seeking treatment at an early intervention outpatient mood and anxiety program. *Psychol Health Med.* 2017;22(8):978-986.

12.	Bandelow B, Michaelis S. Epidemiology of anxiety disorders in the 21st century. *Dialogues Clin Neurosci.* 2015;17(3):327-335.

13.	Association CMH. Children, Youth, and Anxiety. <https://cmha.ca/documents/children-youth-and-anxiety>. Published 2018. Accessed March 27, 2021.

14.	Ontario ADAo. Anxiety and Youth. <http://www.anxietydisordersontario.ca/anxiety-resource-centre/anxiety-and-youth/>. Published 2021. Accessed March 27, 2021.

15.	Centre A. Frequent Questions about Anxiety, Anxiety Disorder, Anxiety Attacks (Panic Attacks), etc. <https://www.anxietycentre.com/anxiety-faq.shtml>. Published 2021. Accessed March 27, 2021.

16.	Health TM. Generalized Anxiety Disorder. <http://mentalhealthliteracy.org/mental-disorders/generalized-anxiety-disorder/>. Published 2021. Accessed March 27, 2021.

17.	InnerHour. The InnerHour Experience. <https://www.theinnerhour.com/>. Accessed April 4, 2021.

18.	Solutions BS. WellMind. <https://play.google.com/store/apps/details?id=com.bluestepsolutions.wellmind&hl=en_CA&gl=US>. Published 2017. Accessed April 4, 2021.

19.	Health VB. Causes, Symptoms & Effects of Anxiety. <https://www.villagebh.com/disorders/anxiety/symptoms-signs-effects/>. Published 2021. Accessed March 27, 2021.

20.	Manual M. Generalized Anxiety Disorder in Children. <https://www.merckmanuals.com/home/children-s-health-issues/mental-health-disorders-in-children-and-adolescents/generalized-anxiety-disorder-in-children>. Published 2019. Accessed March 27, 2021.

21.	Healthdirect. Anxiety in teenagers. <https://www.healthdirect.gov.au/anxiety-in-teenagers>. Published 2020. Accessed March 27, 2021.

22.	service oh. Anxiety disorders in children. <https://www2.hse.ie/conditions/mental-health/anxiety-disorders-in-children.html>. Published 2020. Accessed March 27, 2021.

23.	KidsHealth. Anxiety. <https://www.kidshealth.org.nz/anxiety>. Published 2021. Accessed March 27, 2021.

24.	Aspiro. 13 Signs of Anxiety in Young Adults and Teenagers. <https://aspiroadventure.com/blog/13-signs-of-anxiety-in-teens-and-young-adults/>. Published 2019. Accessed March 27, 2021.

25.	NHS. Anxiety disorders in children. <https://www.nhs.uk/mental-health/children-and-young-adults/advice-for-parents/anxiety-disorders-in-children/>. Published 2020. Accessed March 27, 2021.

26.	Society TCs. Anxiety. <https://www.childrenssociety.org.uk/information/young-people/well-being/resources/anxiety>. Accessed March 27, 2021.

27.	Centre F. Anxiety in Children and Youth. <https://www.famcentre.ca/anxiety-in-children-and-youth/>. Published 2017. Accessed March 27, 2021.

28.	Diagnostics M. Find Out If You Have Anxiety. <https://www.mind-diagnostics.org/anxiety-test?utm_source=AdWords&utm_medium=Search_PPC_c&utm_term=anxiety%20disorder_b&utm_content=88328057960&network=g&placement=&target=&matchtype=b&utm_campaign=6453264419&ad_type=mind-diagnostics&adposition=&gclid=Cj0K>. Published 2021. Accessed March 27, 2021.

29.	Scrandis DA. Anxiety disorders in adolescents. *Nurse Practitioner.* 2019;44(8):12-14.

30.	Spence SH, Zubrick SR, Lawrence D. A profile of social, separation and generalized anxiety disorders in an Australian nationally representative sample of children and adolescents: Prevalence, comorbidity and correlates. *Aust N Z J Psychiatry.* 2018;52(5):446-460.

31.	Centre LHS. Information About Mood & Anxiety. <https://www.lhsc.on.ca/femap-first-episode-mood-and-anxiety-program/information-about-mood-anxiety>. Published 2021. Accessed March 27, 2021.

32.	Centre MUH. Anxious children: Treating a crippling disorder. <https://www.thechildren.com/health-info/conditions-and-illnesses/anxious-children-treating-crippling-disorder>. Accessed March 28, 2021.

33.	Association CMH. Child and Youth Mental Health. <https://ontario.cmha.ca/mental-health/child-and-youth-mental-health/>. Accessed March 28, 2021.

34.	Association CMH. Anxiety Disorders. <https://cmha.bc.ca/documents/anxiety-disorders/>. Published 2013. Accessed March 28, 2021.

35.	TeensHealth. Anxiety Disorders. <https://kidshealth.org/en/teens/anxiety.html?WT.ac=ctg#catmental-health>. Published 2014. Accessed March 28, 2021.

36.	BC F. Anxiety Self-check. <https://foundrybc.ca/quiz/anxiety-self-check/?return_page=1292>. Accessed April 4, 2021.

37.	HeretoHelp. For Youth: Learn about Anxiety. <https://www.heretohelp.bc.ca/infosheet/for-youth-learn-about-anxiety>. Published 2019. Accessed March 28, 2021.

38.	Manitoba HC. Teen Clinic Mental Health Toolkit. <https://www.gov.mb.ca/healthychild/mcad/teen_clinic_mental_health_toolkit.pdf>. Accessed March 28, 2021.

39.	Stem4. Anxiety. <https://stem4.org.uk/anxiety/>. Published 2012. Accessed March 28, 2021.

40.	Headspace. what is anxiety & the effects on mental health. <https://headspace.org.au/young-people/what-is-anxiety-and-the-effects-on-mental-health/>. Published 2021. Accessed March 28, 2021.

41.	Prevention CfDCa. Anxiety and Depression in Children. <https://www.cdc.gov/childrensmentalhealth/depression.html>. Published 2020. Accessed March 28, 2021.

42.	Psychiatry AAoCA. Your Adolescent - Anxiety and Avoidant Disorders. <https://www.aacap.org/AACAP/Families_and_Youth/Resource_Centers/Anxiety_Disorder_Resource_Center/Your_Adolescent_Anxiety_and_Avoidant_Disorders.aspx>. Accessed April 2, 2021.

43.	healthline. Anxiety Diagnosis. <https://www.healthline.com/health/anxiety-diagnosis#outlook>. Published 2021. Accessed April 2, 2021.

44.	Academy N. How to Recognize Anxiety in Teenagers. <https://www.newportacademy.com/resources/mental-health/how-to-recognize-teen-anxiety-disorders/>. Published 2018. Accessed April 2, 2021.

45.	Clinic C. Anxiety Disorders. <https://my.clevelandclinic.org/health/diseases/9536-anxiety-disorders>. Published 2020. Accessed April 2, 2021.

46.	Understood. Signs of Anxiety in Tweens and Teens. <https://www.understood.org/en/friends-feelings/managing-feelings/stress-anxiety/signs-your-teen-or-tween-is-struggling-with-anxiety>. Published 2021. Accessed April 2, 2021.

47.	raisingchildren.net.au. Anxiety disorders in teenagers. <https://raisingchildren.net.au/pre-teens/mental-health-physical-health/stress-anxiety-depression/anxiety-disorders>. Published 2021. Accessed April 2, 2021.

48.	Hospital BCs. Anxiety Disorders Symptoms & Causes. <https://www.childrenshospital.org/conditions-and-treatments/conditions/a/anxiety-disorders/symptoms-and-causes>. Published 2021. Accessed April 2, 2021.

49.	Help B. Anxiety Articles. <https://www.betterhelp.com/advice/anxiety/>. Published 2021. Accessed April 2, 2021.

50.	inform N. Anxiety Disorders in Children. <https://www.nhsinform.scot/illnesses-and-conditions/mental-health/anxiety-disorders-in-children>. Published 2020. Accessed April 2, 2021.

51.	Sinai C. Generalized Anxiety Disorder (GAD) in Children and Teens. <https://www.cedars-sinai.org/health-library/diseases-and-conditions---pediatrics/g/generalized-anxiety-disorder-gad-in-children.html>. Published 2021. Accessed April 2, 2021.

52.	Health F. 10 Common Symptoms of Anxiety Disorder. <https://facty.com/conditions/anxiety-disorder/10-common-symptoms-of-anxiety-disorder/?style=quick&utm_source=adwords->. Published 2020. Accessed April 2, 2021.

53.	Foundation BaBR. Anxiety FAQs. <https://www.bbrfoundation.org/faq/frequently-asked-questions-about-anxiety?gclid=CjwKCAiAnvj9BRA4EiwAuUMDf4uY-2P1fnBqaJJ29vn-VNgfeO3w7zV68OWpkgA0AXqZZjUGjGw-yhoCm1EQAvD_BwE>. Published 2020. Accessed April 2, 2021.

54.	Psycom. 6 Hidden Signs of Teen Anxiety. <https://www.psycom.net/hidden-signs-teen-anxiety/>. Published 2021. Accessed April 2, 2021.

55.	healthychildren.org. Anxiety in Teens in Rising: What's Going On? <https://www.healthychildren.org/English/health-issues/conditions/emotional-problems/Pages/Anxiety-Disorders.aspx>. Published 2021. Accessed April 2, 2021.

56.	Sigmund H. Anxiety in Teens - How to Help a Teenager Deal with Anxiety. <https://www.heysigmund.com/anxiety-in-teens/>. Published 2021. Accessed April 2, 2021.

57.	Center UoRM. Generalized Anxiety Disorder (GAD) in Children and Teens. <https://www.urmc.rochester.edu/encyclopedia/content.aspx?ContentTypeID=90&ContentID=P02565>. Published 2021. Accessed April 2, 2021.

58.	Agers S. Anxiety - What Every Young Person Should Know. <https://www.screenagersmovie.com/tech-talk-tuesdays/anxiety-what-every-young-person-should-know>. Accessed April 2, 2021.

59.	Minds Y. Anxiety. <https://youngminds.org.uk/find-help/conditions/anxiety/>. Published 2021. Accessed April 4.

60.	Centers ET. Generalized Anxiety Disorder in Teens. <https://evolvetreatment.com/parent-guides/anxiety/>. Published 2021. Accessed April 4, 2021.

61.	Van Ameringen M, Simpson W, Patterson B, Turna J. Internet screening for anxiety disorders: Treatment-seeking outcomes in a three-month follow-up study. *Psychiatry Res.* 2015;230(2):689-694.

62.	Moir F, Fernando AT, 3rd, Kumar S, Henning M, Moyes SA, Elley CR. Computer Assisted Learning for the Mind (CALM): the mental health of medical students and their use of a self-help website. *N Z Med J.* 2015;128(1411):51-58.
