## Supplemental Table 2 for "Online anxiety resources for Canadian youth: a systematic environmental scan"

**Table 2. Characteristics of Anxiety Self-Assessment Resources for Use by Youth**

| **Assessment Name**  **Modality**  **Country** | **Organization Type** | **Foundational Tool** | **Next Steps** | **Resources Provided** | **Provides Internal Self-Management Support** | **Provides anxiety score (i.e. number)** | **Provides a statement** | **Provides Descriptive Text** | **Number of Questions Asked** | **Explicitly for Youth** |
| --- | --- | --- | --- | --- | --- | --- | --- | --- | --- | --- |
| Anxiety Test^1^  Application  USA | Private - individual/company | Yes (GAD-7) | Promotes help-seeking  Provides resources | Links  Contact Information | No | Yes | Yes | Yes | 7 | No |
| Link: <https://play.google.com/store/apps/details?id=com.feartools.anxietytest&hl=en_CA&gl=US> | | | | | | | | | | |
| FearTools^2^  Application  USA | Private - individual/company | Yes (GAD-7) | Promotes help-seeking  Provides resources | Links | Yes (within application) | Yes | Yes | Yes | 7 | No |
| Link: <https://www.feartools.com/> | | | | | | | | | | |
| InnerHour: Self-Care Therapy^3^  Application  India | Private - individual/company | None | Provides resources | Links | Yes (within application) | Yes | Yes | Yes | 13 | No |
| Link: <https://www.theinnerhour.com/> | | | | | | | | | | |
| Pocket Mood Tracker^4^  Application  Canada | Private - individual/company | Yes (GAD-7) | None | NA | No | Yes | Yes | No | 7 | No |
| Link: <https://apps.apple.com/us/app/pocket-mood-tracker/id960876692> | | | | | | | | | | |
| Screen for Child Anxiety Related Disorders (SCARED)^5^  Website  USA | Academic | Yes (SCARED) | None | NA | No | Yes | Yes | No | 41 | Yes |
| Link: <https://www.pediatricbipolar.pitt.edu/resources/instruments> | | | | | | | | | | |
| What's My M3^6^  Website  USA | Private - individual/company | None | Promotes help-seeking  Provides resources | Contact Information | No | Yes | Yes | No | 27, with 4 pre-assessment questions unrelated to anxiety score (i.e., age, gender, place of residence) | No |
| Link: <https://whatsmym3.com/> | | | | | | | | | | |
| Canada Mental Health Association - BC Division^7^  Website  Canada | Provincial Government - MH specific | None | Promotes help-seeking  Provides resources | Links  Contact Information | No | Yes | Yes | No | 11 pre-screeing questions (i.e., age, gender, MH history), 15 questions if mininmal or maximal anxiety answers provided | No |
| Link: <https://www.heretohelp.bc.ca/screening/online/?screen=anxiety> | | | | | | | | | | |
| Foundry BC: Anxiety Self-check^8^  Website  Canada | Provincial Government - MH specific | None | Promotes help-seeking  Provides resources | Links  Contact Information  External Self-management Support | No | No | Yes | Yes | 11 | Yes |
| Link: <https://foundrybc.ca/quiz/anxiety-self-check/?return_page=1292> | | | | | | | | | | |
| Kids Help Phone^9^  Website  Canada | Private - foundation/charity | Yes (GAD-7) | Promotes help-seeking  Provides resources | Links  Contact Information | No | No | No | No | 7 | Yes |
| Link: <https://kidshelpphone.ca/get-info/questionnaire-reflecting-on-feelings-of-anxiety/> | | | | | | | | | | |
| NHS: Mood self-assessment^10^  Website  UK | Federal Government - general | None | Promotes help-seeking  Provides resources | Links  Contact Information | No | Yes | No | No | 18 | No |
| Link: <https://www.nhs.uk/conditions/stress-anxiety-depression/mood-self-assessment/> | | | | | | | | | | |
| Anxiety and Depression Society of America: GAD Screening Tool^11^  Website  USA | Federal Government - general | Yes (GAD-IV) | Promotes help-seeking | NA | No | No | No | No | 4 pre-screening questions, 13 questions with option to list stressors | No |
| Link: <https://adaa.org/screening-generalized-anxiety-disorder-gad> | | | | | | | | | | |
| PSYCOM: Anxiety Self-Assessment^12^  Website  USA | Private - individual/company | None | Promotes help-seeking | NA | No | No | Yes | No | 15 | No |
| Link: <https://www.psycom.net/anxiety-test> | | | | | | | | | | |
| Black Dog Institute: Anxiety Self Test^13^  Website  Australia | Private - foundation/charity | None | Promotes help-seeking  Provides resources | Links  Contact Information  External Self-management Support | No | No | Yes | No | 5 | No |
| Link: <https://www.blackdoginstitute.org.au/resources-support/digital-tools-apps/anxiety-self-test/> | | | | | | | | | | |
| Psychology Today: Anxiety Test^14^  Website  USA | Private - individual/company | None | None | NA | No | Yes | Yes | Yes | 42 | No |
| Link: <https://www.psychologytoday.com/ca/tests/health/anxiety-test> | | | | | | | | | | |
| PsychCentral: Anxiety Screening Test^15^  Website  USA | Private - individual/company | Yes (DSM-5) | Promotes help-seeking  Provides resources | Contact Information for internal psychologists | No | Yes | Yes | Yes | 22 | No |
| Link: <https://psychcentral.com/quizzes/anxiety-quiz/> | | | | | | | | | | |
| Anxiety House Brisbane: Self Assessment Quiz^16^  Website  Australia | Private - individual/company | Yes (GAD-7) | Promotes help-seeking from internal psychologists  Provides resources | Contact Information for internal psychologists | No | Yes | Yes | No | 7 | No |
| Link: <https://anxietyhouse.com.au/appointments/self-assessment-quiz/> | | | | | | | | | | |
| Mind Diagnostics^17^  Website  USA | Private - individual/company | Yes (GAD-7) | Promotes help-seeking  Provides resources | Contact Information for internal psychologists | No | Yes | Yes | Yes | 7 | No |
| Link: <https://www.mind-diagnostics.org/anxiety-test?utm_source=AdWords&utm_medium=Search_PPC_c&utm_term=anxiety%20disorder_b&utm_content=88328057960&network=g&placement=&target=&matchtype=b&utm_campaign=6453264419&ad_type=mind-diagnostics&adposition=&gclid=Cj0K> | | | | | | | | | | |

**Abbreviations:** DSM = Diagnostic and Statistical Manual of Mental Disorders; GAD = Generalized Anxiety Disorder; MH = Mental Health

1.	LLC IH. Anxiety Test. <https://play.google.com/store/apps/details?id=com.feartools.anxietytest&hl=en_CA&gl=US>. Published 2020. Accessed April 4, 2021.

2.	LLC IH. FearTools. <https://www.feartools.com/>. Published December 2016. Accessed April 4, 2021.

3.	InnerHour. The InnerHour Experience. <https://www.theinnerhour.com/>. Accessed April 4, 2021.

4.	Taylor J. Pocket Mood Tracker. <https://apps.apple.com/us/app/pocket-mood-tracker/id960876692>. Published 2015. Accessed April 4, 2021.

5.	Pittsburgh Uo. Child and Adolescent Bipolar Spectrum Services. <https://www.pediatricbipolar.pitt.edu/resources/instruments>. Published 2021. Accessed April 4, 2021.

6.	Information M. Take Control of Your Mental Health. <https://whatsmym3.com/>. Published 2018. Accessed April 4, 2021.

7.	HereToHelp. Online Screenings. <https://www.heretohelp.bc.ca/screening/online/?screen=anxiety>. Accessed April 4, 2021.

8.	BC F. Anxiety Self-check. <https://foundrybc.ca/quiz/anxiety-self-check/?return_page=1292>. Accessed April 4, 2021.

9.	Phone KH. Questionnaire: Reflecting on feelings of anxiety. <https://kidshelpphone.ca/get-info/questionnaire-reflecting-on-feelings-of-anxiety/>. Accessed April 4, 2021.

10.	NHS. Depression and anxiety self-assessment quiz. <https://www.nhs.uk/mental-health/self-help/guides-tools-and-activities/depression-anxiety-self-assessment-quiz/>. Published 2020. Accessed April 4, 2021.

11.	America ADAo. Screening for Generalized Anxiety Disorder (GAD). <https://adaa.org/screening-generalized-anxiety-disorder-gad>. Accessed April 4, 2021.

12.	PSYCOM. Anxiety Test (Self-Assessment). <https://www.psycom.net/anxiety-test>. Published 2021. Accessed April 4, 2021.

13.	Institute BD. Anxiety self test. <https://www.blackdoginstitute.org.au/resources-support/digital-tools-apps/anxiety-self-test/>. Published 2021. Accessed April 4, 2021.

14.	Today P. Anxiety Test. <https://www.psychologytoday.com/ca/tests/health/anxiety-test>. Published 2021. Accessed April 4, 2021.

15.	PsychCentral. Anxiety Screening Test. <https://psychcentral.com/quizzes/anxiety-quiz#Learn-More-About-Anxiety>. Accessed April 4, 2021.

16.	Brisbane AH. Self Assessment Quiz. <https://anxietyhouse.com.au/appointments/self-assessment-quiz/>. Published 2017. Accessed April 4, 2021.

17.	Diagnostics M. Find Out If You Have Anxiety. <https://www.mind-diagnostics.org/anxiety-test?utm_source=AdWords&utm_medium=Search_PPC_c&utm_term=anxiety%20disorder_b&utm_content=88328057960&network=g&placement=&target=&matchtype=b&utm_campaign=6453264419&ad_type=mind-diagnostics&adposition=&gclid=Cj0K>. Published 2021. Accessed March 27, 2021.
