## Supplemental Table 3 for "Online anxiety resources for Canadian youth: a systematic environmental scan"

**Table 3. Study Characteristics for Help-seeking related to anxiety among youth**

| **First Author (Year)**  **Country**  **Study Design** | **Data Collection Methods** | **Number of Participants** | **Description of Participants** | **Age: Mean (SD), Range**  **Male (n, %)**  **Those with Anxiety (n, %)** | **Type of Help**  **Help-seeking Experiences, Barriers, and Predictors** | **Description of Information Needs** |
| --- | --- | --- | --- | --- | --- | --- |
| **PROSPECTIVE STUDIES** | | | | | | |
| Moir (2015)^1^  New Zealand  Prospective Cohort | GAD-7 | N = 321 | Medical students in 2^nd^ and 3^rd^ year | 22.2 (2.9) years  154 (48%)  At risk of any degree anxiety = 103 (37%)  Moderate/severe anxiety = 6 (2%) Moderate anxiety = 20 (7%) Mild anxiety = 77 (28%) | Website access of CALM  Predictor: Those who accessed CALM had significantly higher anxiety scores at baseline than those who did not access CALM | NR |
| **MIXED METHODS STUDIES** | | | | | | |
| Elshire (2019)^2^  United States  Mixed methods (dissertation) | 60-minute narratives of athletes. and person-to-person interviews with selected participants | N = 17 | Undergraduate students from NCAA Division institution in Southwest region of the United States | NR  5 (29%)  NR | Mental health support  Experiences: loss of identity  "I think it’s nice that there is someone (sports psychologist) that understands the sports side, but, I would definitely want to be talking about me as a whole and understand that I am stressed about my family, too and other things.”  Barriers: The most common reason for not seeking mental health services was a lack of time. Unfortunately, it might not just be a lack of free time that prevents student-athletes from seeking mental health services, it could prove to be exhaustion. | NR |
| **CROSS-SECTIONAL STUDIES** | | | | | | |
| Calear (2020)^3^  Australia  Cross-sectional | GASS; Attitudes Toward Seeking Professional Psychological Help Scale | N = 1767 | Adolescents from 30 schools across Australia | 14.8 (0.97) years  620 (36.7%)  On average, participants reported low to moderate levels of current generalized anxiety; 467 (71.7%) had a history of anxiety | Attitudes towards help-seeking  Experiences: Participants had low to moderate positive attitudes towards seeking professional help; 26.1% were not likely to seek professional help  Predictors: Significant predictors of attitudes towards professional help-seeking included GAD-7 score, anxiety literacy score, and GAD stigma | NR |
| Leech (2019)^4^  Australia  Cross-sectional | AHSQ with predisposing factors (anonymity, professionally trained staff, trusted source, ability to text, chat facility and interactive features); (DASS-21) | N = 161 | Young adults, 18 to 24 years living in Australia | 20.6 (1.95) years  46 (28.6%)  NR | Service use  Experiences: 35 (44%) described professional face-to-face support as ‘unhelpful’ or ‘extremely unhelpful’; 29 (37%) were neither satisfied nor dissatisfied; information on social media was rated as ‘unhelpful’ for 33 (38%), ‘neutral’ for 29 (33%), and 26 (30%) described as helpful  Barriers: 92 (57%) ‘lack of money’, 53 (32.9%) lack of transportation, 44 (27.3%) low level of satisfaction with quality of care  Predictors: Those who endorsed symptoms of psychological distress were more likely to access professional resources. Informal help-seeking via social media was weakly and significantly related to sex; men were more likely to seek informal support online; Young adults living with direct family were also more likely to access social media (i.e. Facebook) for emotional support | Informal help-seeking through Facebook 68 (42%); Snapchat 27 (17%); and Instagram 21 (13%)  64 (40%) had previously accessed support via a dedicated online counselling service  Considered critical to seeking online help: 155 (96%) ‘trusted source’ of information, 148 (92%) ‘professionally trained staff’, 140 (87%) ‘anonymity’, 131 (81%) ‘the ability to text/type feelings’ |
| Haavik (2019)^5^  Norway  Cross-sectional | Online survey with open-ended and closed questions adapted from a previous study; Mental health literacy was measured using four vignettes describing young people experiences | N = 1249 | purposive sample of upper secondary schools to ensure participation of students from both rural and urban areas, and university preparatory and vocational studies | 17.6 (1.15) years  550 (44%)  NR | Gender differences and perception of barriers  Barriers: Delay in seeking help (90%); cost of treatment; long waiting time and with perceptions related to stigma, such as fear of receiving a diagnosis and fear of being treated differently  Predictors: Significantly more females than males were aware of the youth health station, school nurse and GP; adolescents from university preparatory studies reported greater awareness of school-based educational–psychological services, whereas adolescents from vocational studies reported greater awareness of GP; females were significantly more likely than males to access youth health station or school nurse; adolescents from university preparatory studies reported significantly more use of all services than those from vocational | NR |
| Grist (2018)^6^  United Kingdom  Cross-sectional | HADS online survey; 11 questions were developed to assess smartphone/tablet internet use and habits | N = 775 | All (n = 876) adolescent girls, aged 11–16 years and attending a UK state-funded secondary schools | 11 – 16 years  0 (0%)  A total of 274 (35.4%) girls scored above the cut-off (≥12) for moderate/severe anxiety | Smartphone/app usage  Experiences: Advantages included: Anonymous = 502 (64.8%); Privacy = 503 (64.9%); Availability = 439 (56.5%); Personal = 399 (51.5%)  Disadvantages included: Accuracy (is information true) = 507 (65.4%); Unauthorised access to information = 438 (56.5%)  Barriers: Worries about others seeing the app highlights stigma associated with mental health and the need to maintain privacy.  Predictors: Scoring above cut-off for anxiety according to HADS: Use the internet for mental health purposes = 14 (5.1%) | NR |
| Havinga (2018)^7^  Netherlands  Cross-sectional | Questionnaire | N = 215 | Patients and their children receiving specialized treatment for depressive and/or anxiety disorders | 18.9 (3.4) years  61 (28.4%)  67 (31.2%) with anxiety | Predictors, type of help not specified  Experiences: 91.9% of offspring eventually seek help; 39.6% of offspring, the time to initial help-seeking was more than two years; probability of help-seeking was substantially higher in the first two years after onset  Predictors: gender; age onset; disorder type  Being female and disorder onset in adolescence or adulthood (relative to childhood) predicted a shorter time to initial help-seeking | NR |
| Van Ameringen (2015)^8^  Canada  Cross-sectional | Online use of MACSCREEN and follow-up survey | N = 103 | NR | 32.4 (12.6) years  24 (23.3%)  46 (44.7%) with anxiety | Those who do or do not seek help while using MACSCREEN  Barriers: Why non-seekers did not access care: fear/lack of desire to take medications (57%); not being comfortable discussing anxiety with physician (28%); their anxiety was not severe enough for treatment (28%)  What would need to change in order for them to seek treatment: “It would need to get worse/ interfere more in my daily life” (44%); “I would need to have the financial means to pay for medication or treatment” (42%); “I would need to be convinced that the treatments work” (34%)  Predictors: Total with GAD = 46 (44.7%); treatment seekers with GAD = 31 (58.5%); non-seekers with GAD = 15 (30%) (P<0.01)  Compared to non-seekers, seekers were more likely to meet screening criteria for GAD (P=0.004) | What would you do with information received from MACSCREEN: seek further help from health professional (85.4%); look for more information online (34.0%); talk it over with family member (25.2%) |
| Romanson (2018)^9^  Canada  Cross-sectional | Modified version of the General Help-Seeking Questionnaire | N = 354 | Undergraduate university students (17 to 25 years of age) | 19.94 (1.62) years  53 (15%)  NR | Use of online or other resources  Barriers: barriers experienced by emerging adults in rural settings included: lack of anonymity, social stigma, limited access to or availability of resources; seeking help online, social media, and tele-psychological services represents an alternative means for individuals in rural areas to access help  Predictors: higher levels of attachment anxiety were significantly associated with greater intentions to seek help online by way of posting to anonymous sources of support and searching for information | NR |
| Langley (2017)^10^  United States  Cross-sectional | GAD-7; help-seeking intentions questionnaire | N = 243 | Over the age of 18 and be located within Australia | 25.58 (10.69) years  47 (19.34%)  66 (27%) met criteria for an anxiety disorder based on the GAD-7 cut-off | Therapist  Barriers: I would not be able to afford treatment = 126 (51.9%); I think I can/should work out my own problems rather than talking to a psychologist = 120 (49.4%); I prefer to seek help from family or friends rather than a psychologist = 82 (33.7%); I am not comfortable discussing with a stranger = 76 (31.3%); I fear I would be criticized by others = 62 (25.5%) | NR |
| Coles (2015)^11^  United States  Cross-sectional | Written responses; Mental Health Literacy Questionnaire for Anxiety Disorders | N = 284 | Undergraduate students at a public university in the United States | 19.3 (1.3) years  116 (41%)  NR | College student recommendations for types of help-seeking  Experiences: Lay recommendations for those diagnosed with GAD:  Seek professional help = 128 (45.1%); Lifestyle changes/informal support = 117 (41.2%); Self-help strategies = 12 (4.3%); Do nothing/wait = 22 (7.8%) | NR |
| Clark (2020)^12^  Australia  Cross-sectional | Attitudes Towards Seeking Professional Help Scale; General Help Seeking Questionnaire; | N = 702 | Participants were required to be male and between 12 and 18 years of age | 14.7 (1.39) years  702 (100%)  NR | Stigma and help-seeking  Experiences: 69 (10%) exhibited stigma in response to the non-clinical vignette; 27 (4%) with GAD in response to the clinical  Participants who exhibited stigma in response to the GAD vignette had more negative attitudes towards online help-seeking than those who had not exhibited stigma  Predictors: Participant attitudes towards informal help-seeking were positively associated with intention to seek help from both informal sources of help-seeking but not formal or online sources. Participant attitudes towards online help-seeking were found to correlate with other attitude measures but not with measures of help-seeking intentions | NR |
| Summerhurst (2017)^13^  Canada  Cross-sectional | Survey with 2 questions: #1 - What was most helpful in recovering from your emotional or mental health concerns; #2 - What was most difficult about your recovery from your emotional or mental health concern | N = 283 | Youth with recent onset of primary mood and/or anxiety concerns | 16 – 26 years  90 (31.8%)  NR | Experiences with help-seeking  Experiences: What was most helpful in assisting you to recover from your emotional or mental health concerns: Talking 52 (17 %) and therapy 45 (15 %)  ‘‘The breathing exercises [my psychiatrist] gave me significantly helped my anxiety’’ (female, age 20) | NR |
| Dayan (2018)^14^  United States  Cross-sectional (dissertation) | Background questionnaire and self-report to determine help-seeking behaviour | N = 91 | College students with mental illness, determined by DASS-21 scores | 19.34 (1.64) years  21 (23.1%)  100% | predictors, type of help not specified  Experiences: significant negative correlation between past mental health help-seeking and negative attitude towards mental illness  Predictors: results show no significant change in help-seeking after receiving information/a class talk. The data consistently found that class talk/information regarding mental health help-seeking was ineffective in increasing help-seeking | Findings suggest that ongoing outreach, educational and awareness campaigns may be effective for reducing unmet needs of mental health services |
| Purcell (2015)^15^  Australia  Cross-sectional | GAD-7; Overall Anxiety Severity and Impairment Scale (OASIS) | N = 802 | Youth participants seeking help from one of four headspace clinical services in Melbourne and Sydney, Australia, between January 2011 and August 2012 | 18.3 (3.2) years  449 (33%)  Moderate to high levels of GAD symptoms in the past 2 weeks | Headspace clinical services in Melbourne and Sydney, Australia  Predictors: age and gender  Women scored significantly higher than men on the GAD-7; total scores on the OASIS, which focuses on avoidance behaviours and the extent to which anxiety interferes with functioning in the past week, also differed by gender and age | NR |
| Arcaro (2017)^16^  Canada  Cross-sectional | Spielberger State Anxiety Inventory | N = 548 | NR | 19.2 (2.7) years  208 (38%)  Depression plus anxiety (29%); anxiety alone (16.5%) | Early intervention program for mood and anxiety disorders  Predictors: Gender was significantly related to the method of access into the program: males were more likely than females to self-refer | NR |
| Gandhi (2016)^17^  Canada  Cross-sectional | provincial and national database | NR | 10 and 24 years living in Ontario between 2006 and 2011 | NR  NR  NR | Psychiatric ED use, psychiatric hospitalizations, outpatient psychiatric care visits  Predictors: Anxiety disorders were the most common reason for ED visits, and accounted for the largest absolute increase of 2.2 per 1000 population (P < 0.001) between 2006 and 2011 | NR |
| **QUALITATIVE STUDIES** | | | | | | |
| Clark (2018)^18,19^  Australia  Qualitative | Interviews | N = 29 | Children 7-12 years with anxiety as main problem | 15.17 (1.91) years  29 (100%)  “Clinical” participants (n = 8) were adolescent males that had experienced symptoms of clinical anxiety (either with or without a co-morbid diagnosis of depression) and had contacted a local mental health provider | Chilled Out consists of 8 online lessons for teens to complete independently over 10 weeks  Barriers: ‘barriers to help-seeking’ identified included stigma, limited knowledge-awareness of information on clinical anxiety, effort, and feeling ‘confronted’ by private emotion through help-seeking.  When exploring the theme of stigma, many adolescents articulated views associated with social norms of masculinity; Help-seeking behaviour was conceptualised as ‘weak’ or ‘not macho’, perceived to potentially compromise their social status leaving them vulnerable to stigma; A subtheme associated with themes of masculinity related to concerns that others would dismiss anxiety as not a ‘real’ illness, further increasing the likelihood of the help-seeker being stigmatised as ‘weak’. A number of interviewees felt that this specific fear would cause them to deny experiencing symptoms both to others and themselves | Themes and common elements of information needs:  Options for help need to be highly visible and easily accessible  Schools should increase the information provided on clinical anxiety within school lessons  Information needs a ‘masculine’ tone, with examples using stereotypical ‘manly’ figures  Most adolescent males, parents, and teachers would lack important information on the symptoms and treatment of clinical anxiety |
| Mental Health Commission of Canada (2018)^20^  Canada  [Video vignettes](https://www.youtube.com/playlist?list=PL2NuAPXp8ohbUt1WW0ga4afMYMmRSr7WZ) | Interviews | N = 7 | Emerging adults | NR  1 (14.3%)  100% | Mental health services in general  Experiences: as a child easier to access and there was concern if you missed an appointment, not as much when you are an adult; no connection between school services and community services  Barriers: small community no services available; parents not supportive; transfer of care when moving communities | Communication between schools and community - info on how to facilitate this; service transition for emerging adults |

**Abbreviations**: AHSQ = Actual Help-Seeking Questionnaire; CALM = Computer Assisted Learning for the Mind; DASS – 21: Depression Anxiety Stress Scales; GAD = Generalised Anxiety Disorder; GASS = Generalised Anxiety Stigma Scale; HADS = Hospital Anxiety and Depression Scale; NCAA = National Collegiate Athletic Association; NR = Not Reported

1.	Moir F, Fernando AT, 3rd, Kumar S, Henning M, Moyes SA, Elley CR. Computer Assisted Learning for the Mind (CALM): the mental health of medical students and their use of a self-help website. *N Z Med J.* 2015;128(1411):51-58.

2.	Elshire-Dulle J. The prevalence of and issues associated with the help seeking behavior among college student-athletes. *Dissertation Abstracts International Section A: Humanities and Social Sciences.* 2019;80(8-A(E)):No Pagination Specified.

3.	Calear AL, Batterham PJ, Torok M, McCallum S. Help-seeking attitudes and intentions for generalised anxiety disorder in adolescents: the role of anxiety literacy and stigma. *Eur Child Adolesc Psychiatry.* 2020;16:16.

4.	Leech T, Dorstyn DS, Li W. eMental health service use among Australian youth: a cross-sectional survey framed by Andersen. *Aust Health Rev.* 2019;16:16.

5.	Haavik L, Joa I, Hatloy K, Stain HJ, Langeveld J. Help seeking for mental health problems in an adolescent population: the effect of gender. *J Ment Health.* 2019;28(5):467-474.

6.	Grist R, Cliffe B, Denne M, Croker A, Stallard P. An online survey of young adolescent girls' use of the internet and smartphone apps for mental health support. *BJPsych Open.* 2018;4(4):302-306.

7.	Havinga PJ, Hartman CA, Visser E, et al. Offspring of depressed and anxious patients: Help-seeking after first onset of a mood and/or anxiety disorder. *J Affect Disord.* 2018;227:618-626.

8.	Van Ameringen M, Simpson W, Patterson B, Turna J. Internet screening for anxiety disorders: Treatment-seeking outcomes in a three-month follow-up study. *Psychiatry Res.* 2015;230(2):689-694.

9.	Romanson EE. Online help seeking in emerging adults: The role of attachment style, emotion regulation, and distress disclosure. *Dissertation Abstracts International: Section B: The Sciences and Engineering.* 2018;79(10-B(E)):No Pagination Specified.

10.	Langley EL, Wootton BM, Grieve R. The utility of the health belief model variables in predicting help-seeking intention for anxiety disorders. *Australian Psychologist.* 2018;53(4):291-301.

11.	Coles ME, Coleman SL, Schubert J. College students' recommendations for dealing with anxiety disorders. *International Journal of Mental Health Promotion.* 2015;17(2):68-77.

12.	Clark LH, Hudson JL, Haider T. Anxiety Specific Mental Health Stigma and Help-Seeking in Adolescent Males. *J.* 2020;29(7):1970-1981.

13.	Summerhurst C, Wammes M, Wrath A, Osuch E. Youth Perspectives on the Mental Health Treatment Process: What Helps, What Hinders? *Community Ment Health J.* 2017;53(1):72-78.

14.	Dayan J. Attitude towards mental illness and help seeking behavior among college students:A pre-post design. *Dissertation Abstracts International: Section B: The Sciences and Engineering.* 2019;80(3-B(E)):No Pagination Specified.

15.	Purcell R, Jorm AF, Hickie IB, et al. Demographic and clinical characteristics of young people seeking help at youth mental health services: baseline findings of the Transitions Study. *Early Interv Psychiatry.* 2015;9(6):487-497.

16.	Arcaro J, Summerhurst C, Vingilis E, Wammes M, Osuch E. Presenting concerns of emerging adults seeking treatment at an early intervention outpatient mood and anxiety program. *Psychol Health Med.* 2017;22(8):978-986.

17.	Gandhi S, Chiu M, Lam K, Cairney JC, Guttmann A, Kurdyak P. Mental Health Service Use Among Children and Youth in Ontario: Population-Based Trends Over Time. *Can J Psychiatry.* 2016;61(2):119-124.

18.	Clark LH, Hudson JL, Dunstan DA, Clark GI. Capturing the attitudes of adolescent males' towards computerised mental health help-seeking. *Australian Psychologist.* 2018;53(5):416-426.

19.	Clark LH, Hudson JL, Dunstan DA, Clark GI. Barriers and facilitating factors to help-seeking for symptoms of clinical anxiety in adolescent males. *Aust J Psychol.* 2018;70(3):225-234.

20.	Canada MHCo. Emerging Adults Seek Change in Mental Health Services. <https://www.youtube.com/playlist?list=PL2NuAPXp8ohbUt1WW0ga4afMYMmRSr7WZ>. Published 2018. Accessed March 27, 2021.
